# “hDOS”: An automated hybrid diffuse optical device for real-time non-invasive tissue monitoring—precision and *in vivo* validation

**DOI:** 10.1101/2025.06.03.25328859

**Authors:** Marta Zanoletti, M. Atif Yaqub, Lorenzo Cortese, Mauro Buttafava, Jacqueline Martínez García, Caterina Amendola, Talyta Carteano, Lorenzo Frabasile, Diego Sanoja Garcia, Claudia Nunzia Guadagno, Tijl Houtbeckers, Umut Karadeniz, Michele Lacerenza, Marco Pagliazzi, Shahrzad Parsa, Tessa Wagenaar, Luc Demarteau, Jakub Tomanik, Alberto Tosi, Udo M. Weigel, Sanathana Konugolu Venkata Sekar, Alessandro Torricelli, Davide Contini, Jaume Mesquida, Turgut Durduran

**Affiliations:** ICFO–Institut de Ciències Fotòniques, The Barcelona Institute of Science and Technology, 08860 Castelldefels (Barcelona), Spain; PIONIRS s.r.l., Via Timavo 24, 20124 Milano, Italy; Politecnico di Milano, Dipartimento di Fisica, Piazza Leonardo Da Vinci 32, 20133 Milano, Italy; ASPHALION s.l., Carrer de Tarragona 151-157, 08014 Barcelona, Spain; BioPixS Ltd – Biophotonics Standards, IPIC, Tyndall National Institute, Lee Maltings Complex, T23 HW11 Cork, Ireland; SPLENDO, Marineweg 5, 2241 TX Wassenaar, Netherlands; Hemophotonics s.l., Avinguda Carl Friedrich Gauss 3, 08860 Castelldefels, Barcelona, Spain; Politecnico di Milano, Dipartimento di Elettronica, Informazione e Bioingegneria, Via Giuseppe Ponzio 34, 20133 Milano, Italy; Consiglio Nazionale delle Ricerche, Istituto di Fotonica e Nanotecnologie, Piazza Leonardo Da Vinci 32, 20133 Milano, Italy; Fondazione IRCCS Ca’ Granda Ospedale Maggiore Policlinico, 20122 Milano, Italy; Critical Care Department, Parc TaulÍ Hospital Universitari. Institut D’Investigació i Innovació Parc TaulÍ I3PT, Plaça Torre de l’Aigua, s/n, 08208 Sabadell, Spain; Institució Catalana de Recerca i Estudis Avançats (ICREA), 08015 Barcelona, Spain

**Keywords:** hybrid diffuse optics, time-domain near infrared spectroscopy, diffuse correlation spectroscopy, intensive care, standardization

## Abstract

**Significance:** A new platform/device is presented that advances hybrid diffuse optical monitors closed to clinical practice, bridging the gap between research-grade optical systems and practical bedside applications. Traditional devices often lack automation, multi-parameter functionality, and operator independence, hence, limiting their usability in demanding clinical environments. By offering automation, user-friendly operation, and overcoming the typical limitations of continuous-wave near-infrared spectroscopy, the hybrid diffuse optical platform (hDOS) provides a more accurate and reliable assessment of both oxygenation and perfusion. This innovation is particularly valuable for monitoring critically ill patients, where precise real-time measurements can directly influence patient management and outcomes.

**Aim:** To design, validate, and characterize the platform hDOS that integrates time-domain near-infrared spectroscopy, diffuse correlation spectroscopy, and a pulse oximeter with an automated vascular occlusion test (VOT). The platform aims to support continuous monitoring and the assessment of peripheral microvascular, and metabolic functions in both clinical and field settings.

**Approach:** The validation strategy for the hDOS device follows a comprehensive approach that goes beyond conventional optical performance assessments. Rather than solely verifying fundamental system parameters, the evaluation comprises of real-world usability, operator and patient safety, and clinical implementation. The device’s precision and usability were rigorously tested *in vivo* through test-retest measurements and comparisons with a commercially available device (INVOS 5100C). This was subsequently followed by a seven-month clinical evaluation at Parc Taulí Hospital Universitari.

**Results:** The device underwent extensive validation, accumulating over 200 hours of usage across approximately 150 measurement sessions. The hDOS device exhibited two-fold lower inter-subject and intra-subject variability in baseline tissue oxygen saturation compared to the INVOS 5100C. Furthermore, during a a vascular occlusion challenge, statistically significant differences were observed between the two systems across all extracted parameters. Finally, as a proof of concept, hDOS successfully detected differences in the microvasculature between a general mixed ICU patient cohort (*n* = 100) and a healthy control group (*n* = 37).

**Conclusions:** Overall, hDOS device has performed well in both bench-top and realistic clinical applications on patients *in vivo*. hDOS device provides a unique combination of parameters, available for the first time in a fully automated, self-contained platform.

## 1 Introduction

Near-infrared spectroscopy (NIRS) is a non-invasive optical technique that utilizes light in the range of ∼ 650-950 nm to monitor tissue properties such as local microvascular blood oxygenation (StO_2_) and blood volume. Over the years, NIRS has evolved as a promising tool for assessing tissue perfusion and metabolism, with various implementations designed to overcome technological limitations.

Commercially available NIRS monitors predominantly employ continuous-wave NIRS (CWNIRS), which is cost-effective and widely used in clinical settings. However, CW-NIRS lacks depth discrimination and cannot differentiate between changes in absorption and scattering (i.e., *µ*_a_ and *µ*′_s_). As a result, it provides only relative measurements of hemoglobin concentration. Several adaptations of CW-NIRS, such as spatially resolved spectroscopy (SRS), broadband NIRS, and second derivative NIRS, have been proposed to improve the accuracy of tissue oxygenation indices.^1, 2^ Despite these advancements, CW-NIRS faces challenges related to precision and reproducibility,^3–5^ making comparisons across studies difficult. Consequently, StO_2_ monitors have not achieved routine clinical implementation.

To address these limitations, time-domain NIRS (TD-NIRS) has emerged as an alternative. Unlike CW-NIRS, TD-NIRS enables depth-resolved measurements and allows direct quantification of both absorption and scattering coefficients. This capability enhances the accuracy of assessing tissue oxygenation and perfusion. Historically, TD-NIRS systems were complex, bulky, and expensive, restricting their widespread adoption. However, advancements in laser sources, detection methods, and miniaturization have led to the development of compact and cost-effective TD-NIRS devices.^6–11^ The commercialization of TD-NIRS^7, 12^ has further accelerated its potential for integration into clinical and research applications.

A significant advancement in optical monitoring has been the development of hybrid instruments that combine TD-NIRS with diffuse correlation spectroscopy (DCS). While TD-NIRS provides depth-resolved information on tissue oxygenation by quantifying absorption and scattering properties, DCS measures microvascular blood flow through the analysis of dynamic light scattering.

This integration allows simultaneous measurements of tissue oxygenation and blood perfusion, offering a comprehensive assessment of microvascular function, addressing the limitation of traditional CW-NIRS. Hybrid NIRS-DCS systems have been applied in neuroscience and preclinical research since the early studies on brain monitoring and animal models^13–17^ and it has been increasingly adopted in a wide range of clinical scenarios^18–27^

One prominent use case of NIRS is hemodynamic monitoring in the intensive care unit (ICU). As a non-invasive, bedside tool, NIRS assesses microvascular reactivity and endothelial function in critically ill patients, when combined with a vascular occlusion test (VOT).^28^ NIRS has been applied in ICU settings, including monitoring acute respiratory distress syndrome^29^ and COVID-19,^30, 31^ weaning from mechanical ventilation,^32–37^ and evaluating microvascular reactivity in sepsis and septic shock.^38–49^ Beyond the ICU, NIRS has been utilized in trauma care,^50–54^ surgery,^55–58^ anesthesia,^43, 59–62^ and severe medical conditions.^63–72^ NIRS has been utilized not only in the critically ills but also in advancing our understanding of microvascular function in healthy subjects^73–80^ and athletic performance.^81–85^

In this work, a detailed characterization and validation of the hDOS device^1^ is presented. The hDOS device is a hybrid optical multi-purpose system designed for non-invasive, bedside assessment of microvascular oxygenation and blood perfusion in critically ill patients. The operation and use case of a former version of the hDOS device was previously described in Ref.^86^

### 1.1 Objectives of the device validation

A holistic approach has been opted for in the evaluation of the hDOS device, going beyond the commonly reported “evaluation of basic performances” approach utilized for similar hybrid devices.^87–90^

The typical approach ensures that the basic optical performance metrics are met. In this case, a step further is taken by considering the replication of the systems and their independent operation in clinical settings. This is conceptually illustrated in Fig. 1, where the corresponding tests and measures implemented to address the problems and needs of the end-user are outlined.

**Fig 1.**
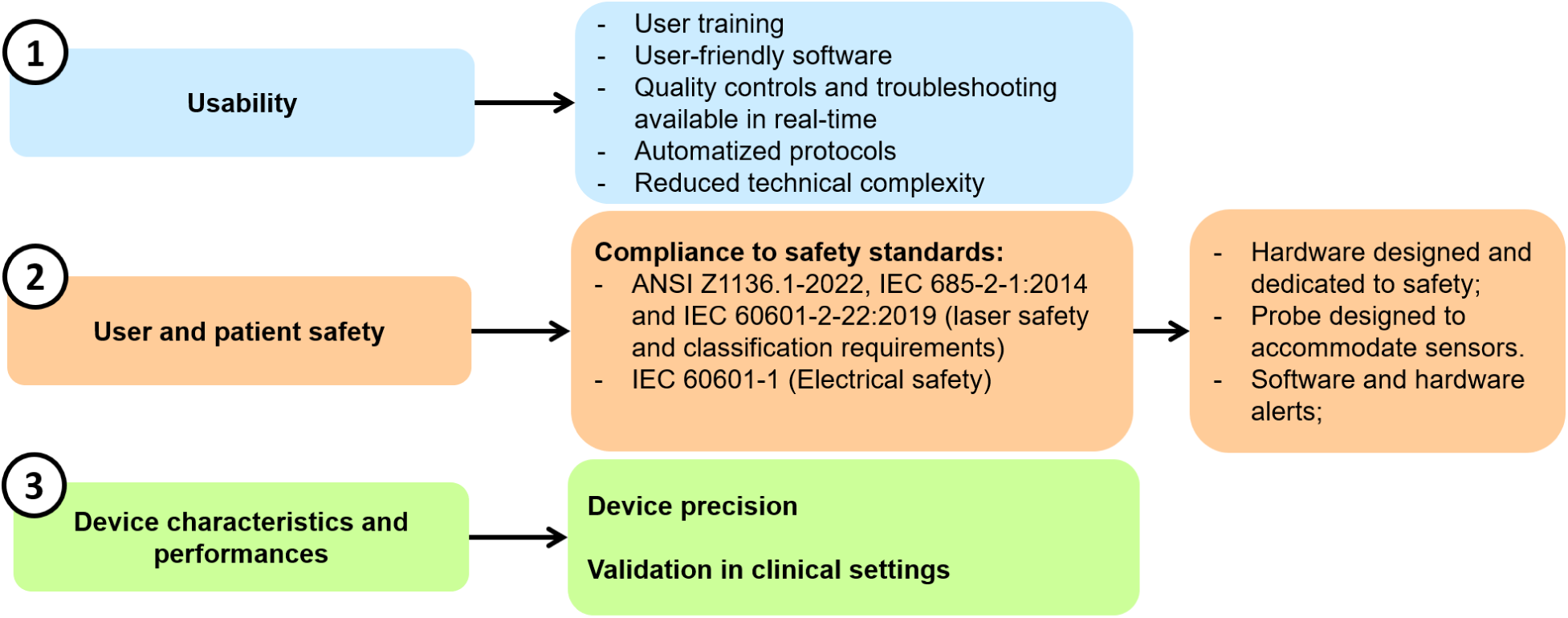
An illustrative overview of the key aspects that has been followed in the deployment of the hDOS device, along with the corresponding measures implemented.

Detailed descriptions of how these measures were integrated during the design phase are provided in section 2.1 and their use in a typical protocol is described in Ref.^86^

Three key aspects have been addressed in the design and development of the hDOS device: (i) usability, (ii) user and patient safety, and (iii) device characteristics and performances.

A typical research system developed by us and other laboratories^19, 87, 88, 91–94^ rely on extensive training and the experience of the operator for clinical study deployment. Here, the system was provided with user-friendly software for independent clinical use with minimal training. Extensive automated safety checks were implemented, detailed data and probe placement metrics were used to provide front panel and on-screen alerts, and software messages were designed to guide users in understanding the signal, identifying problems, and troubleshooting. The software guides users through the protocol and provides real-time access to both raw and processed data, thereby reducing the complexity of operating the device.

Real-time safety measures for both operators and patients have been incorporated through hardware and software tools. Continuous monitoring and quick-response mechanisms are used to maintain safe operation. Quality indicators are also included to alert when the laser is active, environmental light interferes with the signal, or real-time data fitting is not optimal as described in Section 2.

The device has been deployed in the intensive care unit at Parc Taulí Hospital Universitari for seven months. This evaluation was conducted not only for usability and training purposes but also to assess the device performance in an the ICU environment. Tests have been conducted on both tissue-mimicking phantoms and *in vivo*, in order to test the device precision and to set the basic performances in clinical settings. In particular, the device performance during probe repositioning (test-retest) and its susceptibility to interference affecting the two optical modules (as detailed in the dedicated section) were evaluated, and comparisons were made with a commercially available NIRS device used in the clinics.

## 2 Methods

### 2.1 Platform description

This platform was developed in a close-knit collaboration in a modular fashion based on the experience accumulated by several partners involved in this project and is illustrated in Fig. 2. Here we highlight the primary links between the modules and their communication paths to ensure proper operation and include a picture of the device and its accessories in Fig. 3.^86^

**Fig 2.**
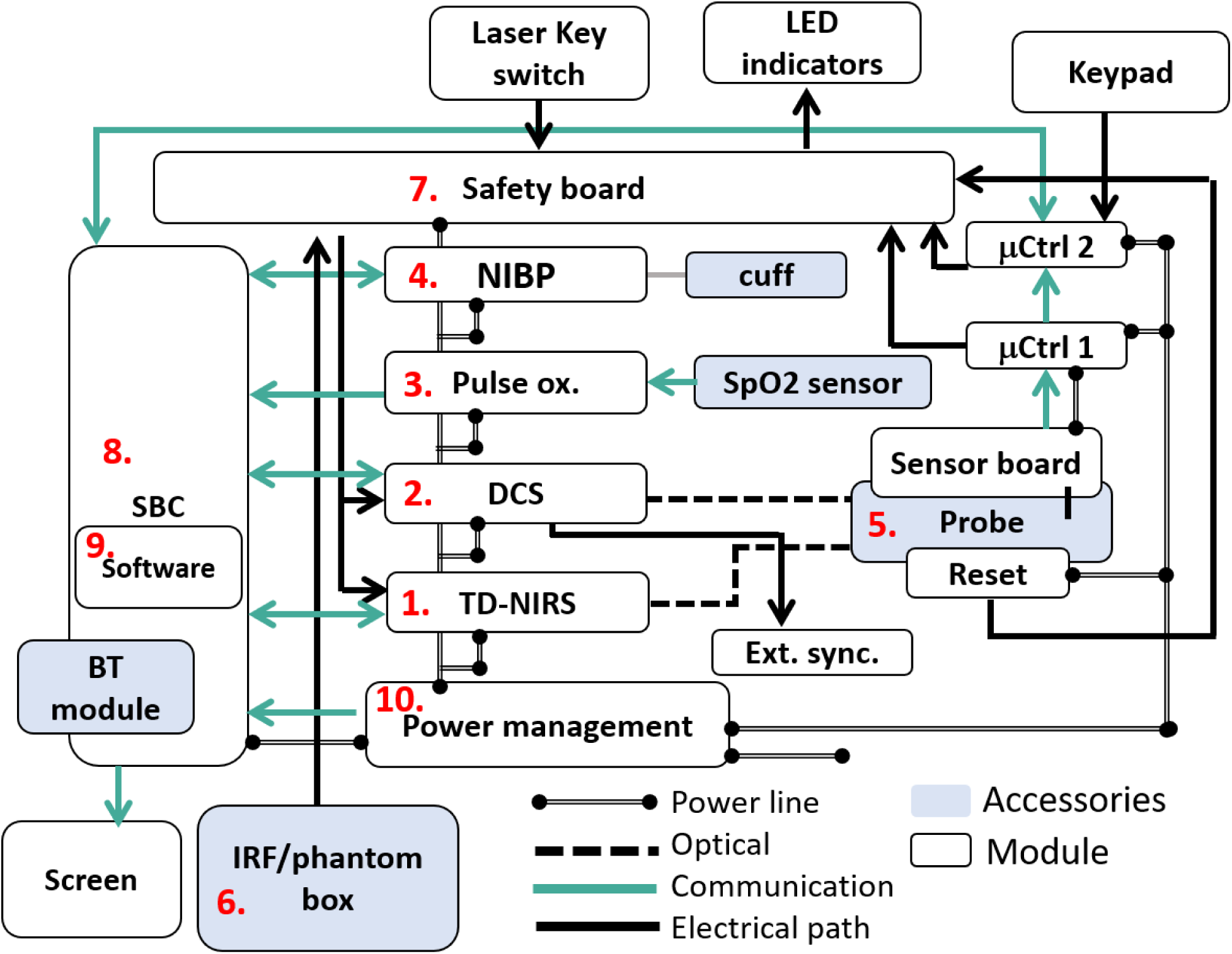
Schematic of the device. 1) TD-NIRS module; 2) DCS module; 3) Pulse oximeter module connected to the SpO_2_ sensor; 4) noninvasive blood pressure module (NIBP) connected to the properly sized cuff; 5) optical probe and sensor board communicating with two microcontrollers (*µ*Ctrl 1 and 2); 6) IRF/phantom box; 7) safety board and primary connection with the other primary modules; 8) single board computer with 9) in-house made software running onboard. A Bluetooth module is available on board for remote control. The SBC is also connected with an internal and/or external screen; 10) power management system. TD-NIRS: time domain near-infrared spectroscopy; DCS: diffuse correlation spectroscopy; Pulse ox.: pulse oximeter; 4. NIBP: noninvasive blood pressure; *µ*Ctrl 1 and 2: microcontrollers 1 and 2; IRF: instrument response function; SBC: single board computer (SBC).

**Fig 3.**
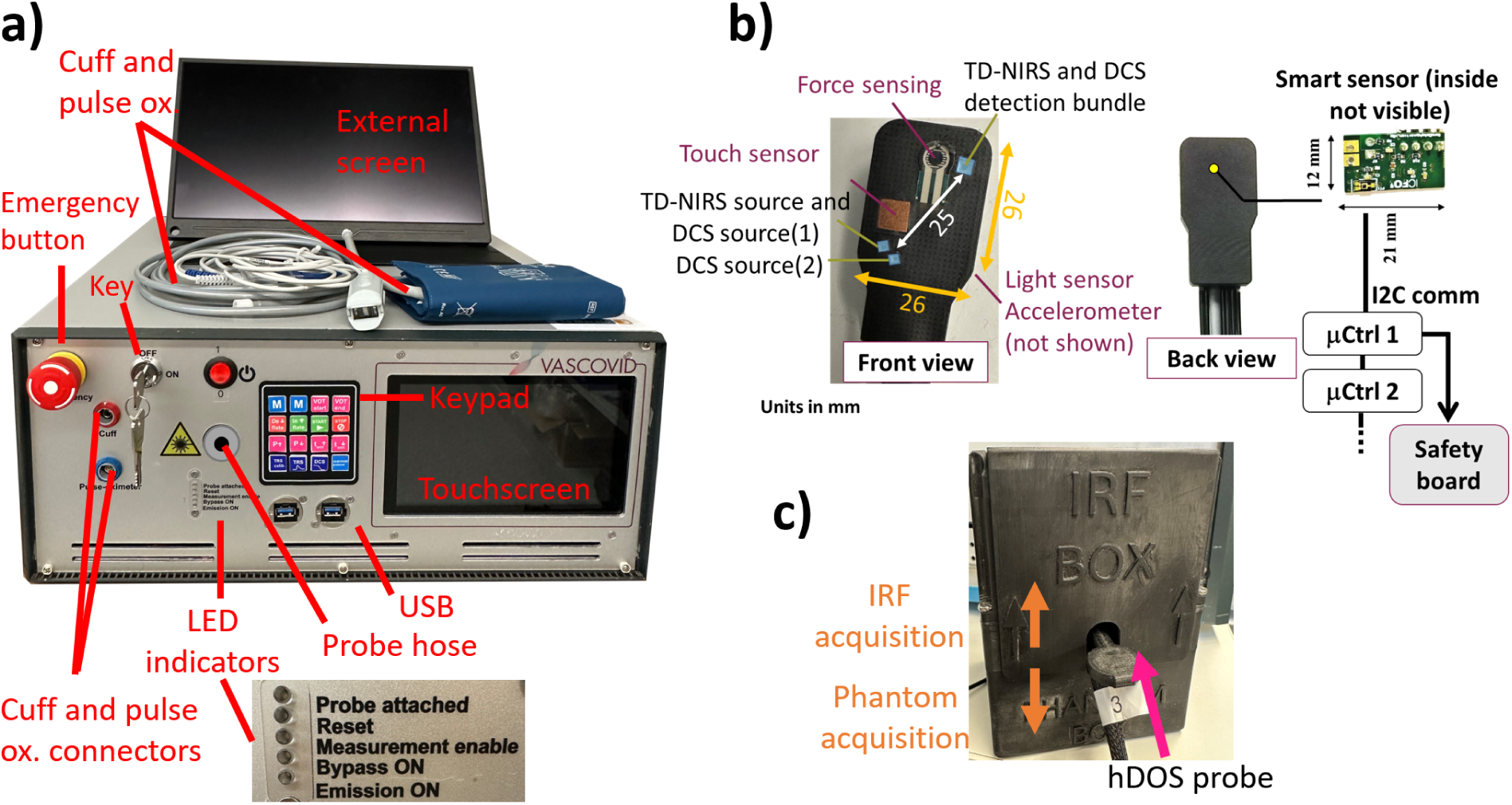
Picture of the hDOS device with its accessories; a) whole platform with main components and a zoom on the LED indicators; b) front and back views of the optical probe together with its smart sensor board embedded within the probe; c) IRF/phantom box.

The following paragraphs will detail each module as indicated in the figure, explaining the design choices and features. A thorough description of the use of a previous (yet very similar) version of this device has been published elsewhere.^86^ Here, we focus on salient technical details and features that define the hDOS device.

**1. Time-domain NIRS, TD-NIRS.** The TD-NIRS module (1) is an original equipment manufacturer (OEM) customized version of the NIRSBOX (PIONIRS s.r.l., Italy).^95^ TD-NIRS^6, 96, 97^ employs two pulsed lasers working at 53 MHz, emitting short pulses at 685 and 828 nm (with the duration in the order of ≈100 ps) that are shone into the tissue by means of a bifurcated bundle composed of two 100/140 *µ*m glass graded index multimode fibers (NA=0.22). The lasers are also coupled with optical attenuators which are commanded via software in order to reach the correct signal-to-noise-ratio (SNR). Laser pulses from each wavelength are injected alternatively into the bundle.

The maximum power injected into the tissue is 3.5 and 3.8 mW for 685 and 828 nm, respectively and it is limited by the maximum permissible emission (MPE) by the ANSI Z1136.1-2022 and IEC 6825-2-1:2014.^98, 99^

Furthermore, the TD-NIRS module is equipped with an interlock system that receive a transistor-to-transistor (TTL) signal that originates from the safety board (8) and it allows for controlling the TD-NIRS laser emission. Diffuse light is then recollected by 1 mm graded index core plastic multimode fiber (NA=0.39) placed at 2.5 cm from the source, which is in turn fed into a silicon photomultiplier detector. Wavelength selection is made by a custom-made dual pass band filter. Both source and detector fibres are hosted in the optical probe (5).

The distribution of time of flight (DTOF) of photons is reconstructed at each wavelength every second, with an integration time of 500 ms, using a custom digital single photon counting unit.

The TD-NIRS module communicates with a single board computer (SBC) (8) to exchange data and time-stamps, as well as other important information such as status from the lasers and other internal controls. Primary communication is via USB 2.0. All the information saved and exchanged with this module are summarized in the Table 5, available in the Appendix.

**2. Diffuse correlation spectroscopy, DCS.** DCS (custom developed based on ICFO/HemoPhotonics S.L., Spain modules)^13, 100, 101^ (2) utilizes long-coherence length (*>*10 m), CW light at 785 nm to quantify the statistics of the diffuse laser speckles as impressed by the movement of red blood cells.^100, 102^

Light at 785 nm is coupled into a fibre splitter (200 *µ*m core and NA = 0.22) and then fed into two step index 400 *µ*m core (NA=0.39) fibers. For this particular case, CW light at 785 nm is limited to 28 mW by the ANSI Z1136.1-2022 and IEC 6825-2-1:2014.^98, 99^ For this reason, as already proposed in other approaches,^103^ the injection point is split into two injection points, which are located on a arc circumference of 3.5 mm with a radius of 2.5 cm, corresponding to the source detector distance for DCS. This approach allows for doubling the SNR of the measurement. This allows to probe still the same volume while accounting for thermal relaxation. CW light is then collected by two 4.4/125 *µ*m core/cladding diameter single mode (NA = 0.13) fibers. Since TD-NIRS and DCS measurements are performed simultaneously, each DCS detection fiber is connected to pigtailed bandpass filter (OZ optics Ltd., Canada).

Finally, the detection fibers are fed each into a single photon avalanche photo diode whose TTL output is utilised by an hardware correlator to construct the intensity autocorrelation function g_2_(*τ*)= *< I*(*t*) · *I*(*t* + *τ*) *> / < I*(*τ*) *>*^2^, where t is the measurement time, *τ* is the lag time and *<* · *>* denotes a time average. In this case, DCS quantifies the g_2_ over an averaging time of 26 ms, allowing the resolution of the pulsatility of blood flow due to the cardiac cycle.

DCS module also allows for a precise synchronization with the TD-NIRS (1) module by an external trigger, a TTL signal (from 1 to 10 Hz), originating from the hardware that calculates the autocorrelation functions. A replica of this signal is available for external connections to eventually synchronize with other clinical monitors and/or data aggregation platforms (Ext. sync.).

Finally, the DCS module communicates to the SBC (8) via USB 2.0, to exchange data collected (g_2_, intensity and time stamps). All the information saved and exchanged with this module are summarized in the Table 6, available in the Appendix.

**3. Pulse-oximeter, SpO_2_ module.** The platform features a pulse oximeter (Medlab GmbH, Germany) (3) that continuously and synchronously calculates the photoplethysmograph signal at 50 Hz (PPG), arterial oxygenation (SpO_2_), heart rate (HR), and perfusion index at 1 Hz. The module communicates with the SBC (8) via serial communication RS-232. All the information saved and exchanged with this module are summarized in the Table 7 and 8 available in the Appendix.

**4. Automated tourniquet, NIBP module.** The automated tourniquet (custom-developed by Medlab GmbH, Germany) (4) has been modified to support rapid inflation and deflation rates, recommended for VOT. Cuff pressure data is transmitted to the SBC (8) at 5 Hz, with a maximum recommended inflation pressure of 300 mmHg. The device includes a range of cuff sizes (Medlab GmbH, Germany) designed to accommodate different limb circumferences. These cuffs are fabricated from biocompatible flexible polyurethane and can be reused following disinfection.

This module communicate with the computer via RS232 serial communication. All the information saved and exchanged with this module are summarized in the Table 7 available in the Appendix.

**5. Probe.** The primary objectives in designing the multimodal probe were to ensure comfort, safety, and data quality. In this paragraph, the rationale and methods behind achieving these goals will be explained. A picture of the probe is shown in Fig. 3 b).

TD-NIRS and DCS fibers are encased in a durable, 3D-printed black rubber material with a shore hardness rating of 85A. The fibers in the probe head are organized such that the sources, including the bifurcated bundle of TD-NIRS and one DCS source, are grouped together and coupled to the tissue using two right-angle 2 mm prisms. For detection, all the TD-NIRS and DCS fibers are combined into a single bundle, which is then coupled with the tissue by using a right-angle 3 mm prism. Each module source and detector pairs are 2.5 cm far from each other. Due to the geometrical arrangement, filters have been positioned in front of the TD-NIRS and DCS to accurately select the wavelengths of interest, as explained in the previous sections.

Each fiber is enclosed in a lightweight vinyl sleeve with a minimal diameter and all are ultimately protected by a meshed sleeve.

The arrangement of the source and detector is designed to accommodate a touch sensor (7 x 7 mm x 0.02 inch copper plate) for detecting contact, as well as a force sensing resistor (FSR400) for monitoring pressure changes exerted by the probe on the tissue.

Both capacitive and force sensor terminations are connected through coaxial cables to their respective sensing chip and feedback circuit embedded on a custom printed circuit board placed on the probe head.

In addition to a contact and force sensor, a 3-axis accelerometer and a photodiode have been included.

On the back of the probe head, a smart sensor board (12 x 21 mm) is enclosed, which transmits data to a microcontroller (*µ*Ctrl 1) via I2C communication protocols. The *µ*Ctrl 1, based on a pre-set threshold on the touch sensor, sends a TTL signal to the safety board (7) to indicate the probe attachment to the tissue. The *µ*Ctrl 1 also transmits data to a second microcontroller (*µ*Ctrl 2), which then communicates with the SBC (8). The data received from *µ*Ctrl 2 are utilized by the software to set user and device alerts. The use of two sequential microcontrollers is aimed at minimizing the possibility of accessing the sensor board and overriding the safety signals.

Due to the presence of signal and power lines, an electrical isolator is positioned along the probe to decouple the patient from the device. Additionally, a reset switch has been implemented, which must be pressed if probe detachment is detected for more than 10 s, in compliance with the latest safety standard IEC 60601-2-22:2019.^104^ If the switch is not pressed laser emission is not enabled. Collectively, these and other measures (see below) taken allow this device to be characterized as being in Laser Class 1C as opposed to Class 3B.

Alerts and indicators of signal quality are available to the user and for post processing as shown in Table 7, available in the Appendix.

**6. IRF/phantom box.** The platform is equipped with a smart box that facilitates daily instrument response function (IRF) assessment^105^ which also includes a durable, solid tissue-simulating phantom (BioPixS Limited, Ireland) for TD-NIRS quality control. The smart IRF/phantom box (Fig. 3 c) is equipped with an electrical circuit made of strategically placed mechanical switches. When the probe is inserted into the box, these switches are activated, generating a signal that is fed to the on-board safety control unit that in turn alerts the user by activating the “probe attached” LED indicator on the front panel and engages the internal interlocks for the TD-NIRS and DCS lasers. This feature is designed to simulate the attachment to tissue via a contact sensor, which does not function with the phantom. Once the “attachment” is detected, the operator is allowed to acquire an IRF and/or a phantom measurement. The IRF measurement involves acquiring a single DTOF for 1 s, aiming for a photon count rate of 10^6^ photons per second for each wavelength. The key IRF parameters, such as the temporal position of its barycenter and full-width-half-maximum (FWHM), are calculated real time by the on board software (9). In particular, the FWHM represents the temporal resolution, while its barycenter reflects the stability of the laser, detection, and acquisition chain, which ultimately impacts the retrieval of the tissue’s optical properties.

The phantom utilized in this case, is made of silicone background, with absorption contribution given by carbon black and scattering given by titanium dioxide, with desired optical properties at 685 nm *µ*_a_ = 0.23 cm^−1^ (*µ*′_s_ =12.8 cm^−1^) and at 830 nm *µ*_a_ = 0.19 cm^−1^ (*µ*′_s_ = 9.5 cm^−1^) (Biopixs, s.l., Ireland). The phantom measurement in particular is made of 20 repetitions with acquisition time of 1 s and target count rate of 10^6^ counts per second per wavelength. Also, for the phantom measurements the on board software (9) calculates in real time the FWHM, the barycenter as well as the optical properties of the phantom.

Throughout the entire validation process, all users, including lab technicians and clinicians, have been guided to acquire an IRF to accurately calibrate TD-NIRS measurements as well as a phantom measurement at each session. The purpose is to assess the day-to-day performance in terms of precision and reproducibility, as well as quantifying potential degradation of the module over time.

**7. Safety board.** In order to integrate and manage all the safety features from the previously described modules, a dedicated safety board featuring key components such as *µ*Ctrl 1 and 2 has been implemented. The safety board operates by generating the laser-on signal for both TD-NIRS and DCS based on several conditions:

- the key on the front panel must be in the “on” position;
- the user reset button on the probe must be “off”, indicating that the probe is attached and no detachment (monitored by *µ*Ctrl 1) longer than 10 s has occurred;
- *µ*Ctrl 2 must receive the software command to enable the lasers for both systems, subsequently communicating this to the safety board;
- while using the IRF/phantom box the probe must be inserted properly for the “attachement” to be sensed.

If any of these conditions is not fulfilled, the safety board activates/deactivates LEDs placed on the front panel to indicate the necessary action that the user needs to take. This ensures prompt attention to maintain operational safety.

**8. Single board computer, SBC.** The platform is controlled by an industrial-grade single board computer (SBC-230D N4200, ASRock Industrial Computer Corp. Taiwan, R.O.C.), which includes a built-in touchscreen and keypad. The keypad is programmed for performing manual markings, initiating pre-programmed protocols, and managing initial calibration or self-test procedures. It connects to the *µ*Ctrl 2, which communicates any keypress to the SBC. The SBC handles critical functions such as running the software, managing communication with all modules for sending commands and receiving data, and ensuring reliable data storage.

Additionally, remote operation is enabled through Bluetooth technology. This feature is particularly useful in settings like infectious disease triage.

**9. Software.** The onboard software is tailored to meet the needs of medical teams. It provides real-time plots of oxy-, deoxy- and total hemoglobin (HbO, HbR and tHb) concentrations, StO_2_, blood flow index (BFI) and quality parameters for TD-NIRS, DCS, and pulse oximeter readings. How these parameters are extracted in real time has been already described in the Ref.^86^

Operators can also access a device monitor displaying intensity autocorrelation functions for DCS and DTOFs for the TD-NIRS module.

The software suite comprises two applications: one developed in NetBeans IDE 8.2 using Java Development Kit 8 and another in Python 3.11 for Bluetooth communication. Both applications run on the Ubuntu 22.04 LTS operating system. The hDOS device software’s main panel displays time traces and measurements in real-time (Fig. 4 a)). Users can also monitor safety and quality indicators, such as emission and probe attachment status, to ensure optimal device performance (Fig. 4 c)). Different actions are available to the operator. Details about the software and the different protocol available are already explained elsewhere alongside with their functions.^86^

**Fig 4.**
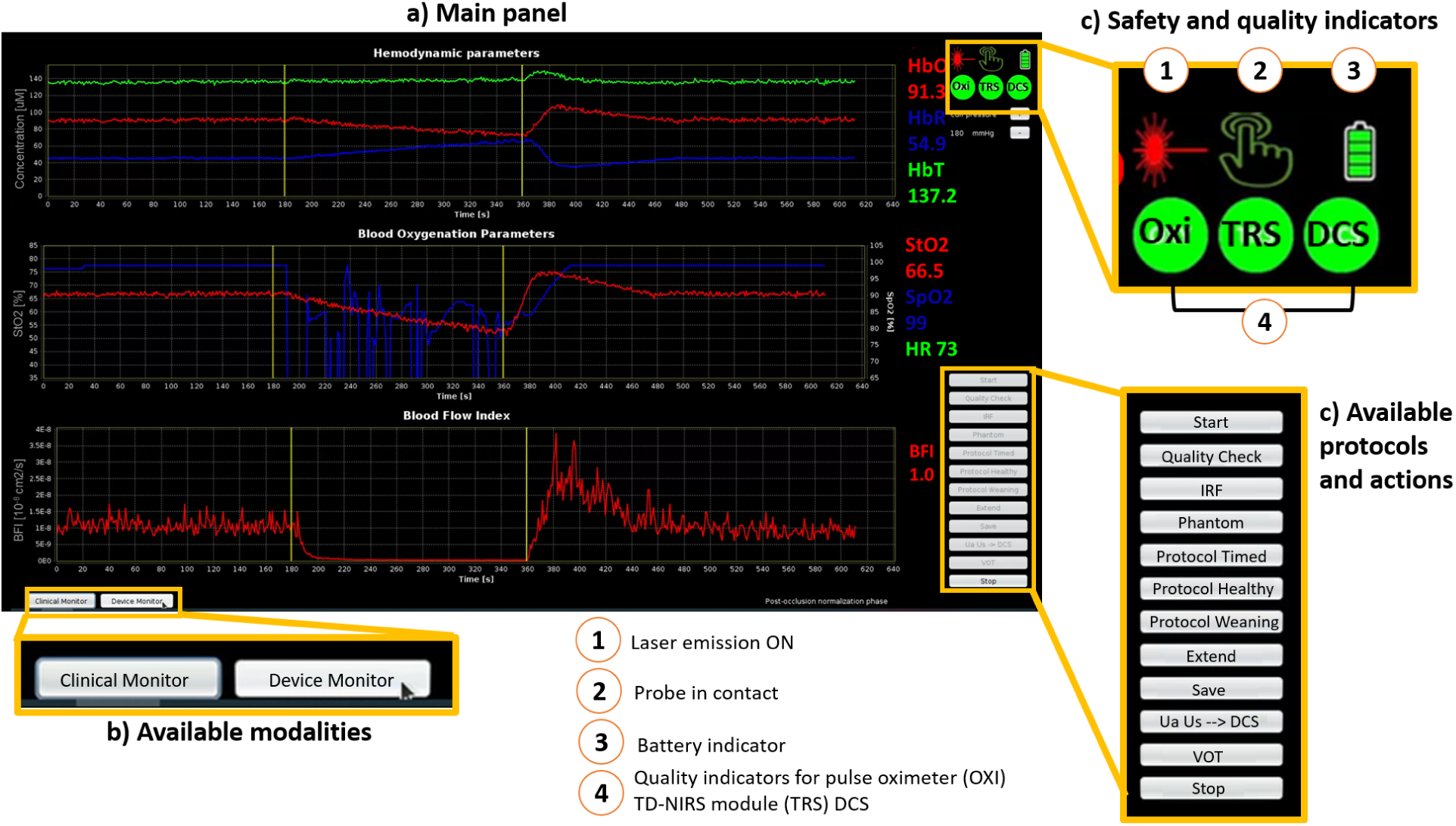
Snapshot of the hDOS device user interface: a) The main panel displays real-time parameters such as hemoglobin concentrations (HbO, HbR, and tHb in *µ*M), StO_2_, SpO_2_ (in %), and BFI (in cm^2^/s; b) The available modalities include a clinical monitor (main panel) for parameter display and a device monitor for raw data quality assessment; c) Real-time safety and quality indicators for SpO_2_ (OXI), TD-NIRS, and DCS (labeled as TRS and DCS, respectively) are provided. A battery indicator is also included since the device operates on battery power; d) available protocols. HbO: oxygenated hemoglobin; HbR: deoxygenated hemoglobin; tHb: total hemoglobin; StO_2_: tissue oxygen saturation; SpO_2_: peripheral oxygen saturation and BFI: blood flow index.

**10. Power management system.** The power management is handled by a dedicated board designed to convert and distribute the necessary power from four battery packs, each connected to its respective power management system (RRC Power Solutions GmbH, Germany). These power management systems can seamlessly switch between battery and external power supply. Additionally, the batteries communicate their charge status and other critical information to *µ*Ctrl 2, ensuring efficient power monitoring and management. The addition of a power management module ensures the device’s usability in challenging settings, such as ICU, where numerous monitors are used simultaneously, and power plugs are not always readily available.

Finally, the device has been tested by means of an electrical safety analyzer which is compliant with the IEC 6060-1 standard.^106^ An emergency button has been incorporated and strategically placed to be easily accessible on the front panel, allowing for a quick action. This safety feature ensures that all the electrical lines are disconnected, thereby stopping the device operation and preventing potential harm to operator and patients.

### 2.2 Analysis method for TD-NIRS and DCS and VOT derived biomarkers

The retrieval of the parameters of interest in real time are explained in Ref.^86^ On the other hand, the parameters derived from the VOT challenge are obtained *a posteriori* with a semi-automated script developed in MATLAB2021b (The MathWorks Inc., Natick, Massachusetts, USA).

#### VOT-derived parameter calculation

The Fig. 5 reports a schematic of the typical response to an ischemic challenge for the StO_2_ and BFI parameters. The VOT corresponds to an initial three minute baseline, a three minute arterial occlusion to a pressure exceeding the systolic pressure by 50 mmHg, and a recovery period of five minutes upon cuff release.

**Fig 5.**
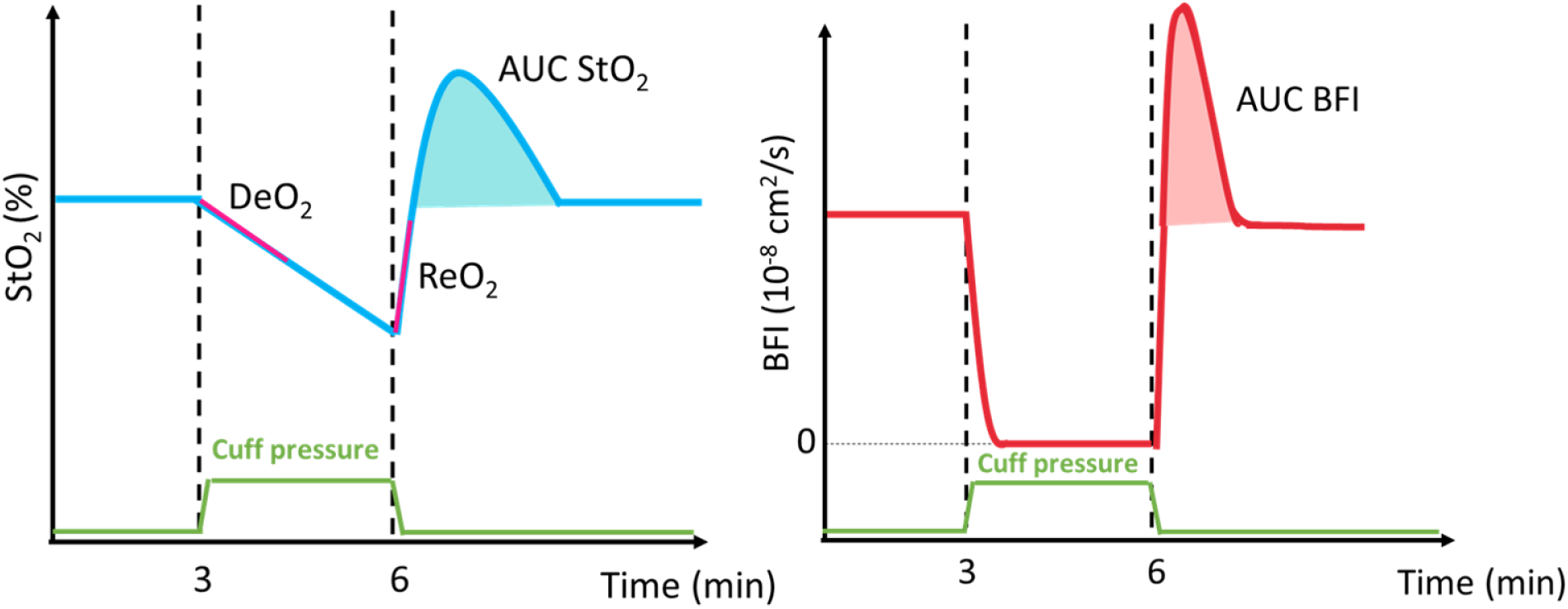
Schematic response of a VOT for StO_2_ (%) and BFI (cm^2^/s). Black vertical dashed lines represent the inflation and the deflation points. The purple lines represent i) the linear fitting of the first minute of occlusion with its slope being the DeO_2_ (%/min) and ii) the linear fitting upon the cuff release, with its slope being the ReO_2_ (%/s). The shaded areas represent under the curve in both StO_2_ (%·min) and BFI (cm^2^).

The VOT-derived parameters are also highlighted: i) baseline phase for both StO_2_ and BFI, ii) deoxygenation rate (DeO_2_) corresponds to the slope of the linear fitting of the first minute of occlusion, iii) reoxygenation rate (ReO_2_) is the slope of the linear fitting from the deflation point (corresponding to the minimum StO_2_ reached during the occlusion) to the intersect of StO_2_ with the second baseline reached in the recovery period and iv) the area under the curve (AUC) StO_2_ and AUC BFI is the area calculated from the first to the second intersect of StO_2_ with the baseline reached in the recovery period. Starting of the measurement, inflation and deflation points correspond to the black lines.

#### Baseline metabolism assessment

The index of local metabolic rate of oxygen extraction for the skeletal muscle (MMRO_2_) is derived at the baseline by the simultaneous and independent assessment of BFI (oxygen transport to the tissue) along with arterial and microvascular tissue oxygenation (oxygen utilization), without the need for a vascular occlusion. In a steady-state condition, MMRO_2_ can be calculated as MMRO_2_ ≈[Hb]·(StO_2_-SpO_2_)/*γ*SpO_2_· BFI, as derived from the Fick’s law,^13, 16, 107^ where [Hb] is the hemoglobin concentration in g/dL and *γ* is the percentage of blood content in the venous compartment.

### 2.3 Precision of the hDOS device

The hDOS device validation has been performed mainly with a focus on the optical modules. The accessory modules, such as the pulse oximeter and the automatized cuff are tested separately by the original equipment manufacturer according to their standard operating procedures. This section focuses on the tests conducted to evaluate the precision of the device, specifically in four areas: (i) variability of key parameters *in vivo* and on phantom at module level; (ii) the variability observed during probe placement and repositioning, and (iii) the device variability under different detected light levels and finally, (iv) optical interferences during measurements.

The following paragraphs will provide an in-depth explanation of the protocols used to address these aspects.

#### 2.3.1 Variability of key parameters at module level

As explained in Section 2.1, point **6**, all users were instructed to acquire a IRF and a phantom measurement prior each session. For each phantom the effective tHb^ph^ and StO_2_^ph^ were computed, using the hemoglobin absorption coefficients at 685 and 828 nm, similar to a previous study.^103^

The coefficient of variation (CV) is a typical figure of merit for the assessment of day-by-day variability and it is defined as the ratio between standard deviation(x) and the mean(x) (in %) over the entire campaign, where x is the measurand of interest, such as FWHM, barycenter, optical properties and effective tHb^ph^ and StO_2_^ph^. In order to simplify the calibration process and improves the overall efficiency of the system, as a proof-of-concept, the first IRF acquired in month 3 was utilized to process all the phantom measurements acquired in that same month. It has been then compared to the standard day-by-day calibration. This comparison was performed using the Wilcoxon signed-rank test with a significance level set at p*<*0.05.

Regarding DCS, it is not possible to monitor in a robust manner quality parameters by utilizing a solid phantom since being static, only contribute to fixed scattering and do not mimic the dynamic changes essential for evaluating flow or motion metrics. A decision was taken to not measure a liquid phantom at each session to ensure a smoother measurement process.

On the other hand, the raw g_2_ is carefully analyzed at each *in vivo* measurement session. The count rate and the *β* can be used as quality parameters. The *β* parameter is a constant number related to the number of modes detected. In a typical DCS device, *β* ≈0.5 and it has to remain stable throughout the whole measurement time. A drop in this parameter might be due to instabilities of the laser, non-coherent light leakage or probe detachment. To test the variability of the DCS module, the CV over the count rate and the *β* are reported during the baseline. For *β* the median and the standard deviation are calculated from 60 s of baseline measurements where the raw g_2_ is integrated by 10 s. In the same 60 s of baseline, the median and standard deviation of the count rate is obtained in kHz. In order to determing if specific trends are visibile, Spearman’s correlation analyses were conducted for both *β* and count rate. Additionally, constant user feedback helped us in identifying fiber issues and power losses, which were promptly addressed.

#### 2.3.2 Test-retest variability

In order to assess the variability due to probe repositioning, a test-retest experiment has been performed on the *brachioradialis* muscle of one subject by repositioning the probe five times and acquiring data for about 200 s at 1 Hz for both DCS and TD-NIRS, similar to what is reported in other works by us and others.^95, 103^ Finally, the mean values of StO_2_ as well as BFI in the five tests and as well as the intra-test and inter-test variability as CV, have been calculated.

#### 2.3.3 Dependence of precision with respect to absolute values

A custom protocol was implemented to assess the device precision and identify small but meaningful alterations in both StO_2_ and BFI. This was done by occluding arterial blood flow at increments of 40%, 60%, 80%, and 100% of the limb occlusion pressure (LOP) during both inflation and deflation. The LOP was identified as the point at which the PPG signal vanishes during limb occlusion. For each occlusion interval, lasting 60 s, the CV was calculated for both BFI and StO_2_, over the last 15 s of the inflation or deflation interval.

The protocol is shown in Figure 6. The dependence of the CV on the mean absolute values calculated across the different inflation/deflation periods was then analyzed, along with how CV changes at the various signal levels, as indicated by the count rate (for DCS) or dynamic range (for TD-NIRS), by means of Spearman’s correlation. The dynamic range is calculated as the ratio as the maximum of the DTOF (in number of photons) over the standard deviation of the background.

**Fig 6.**
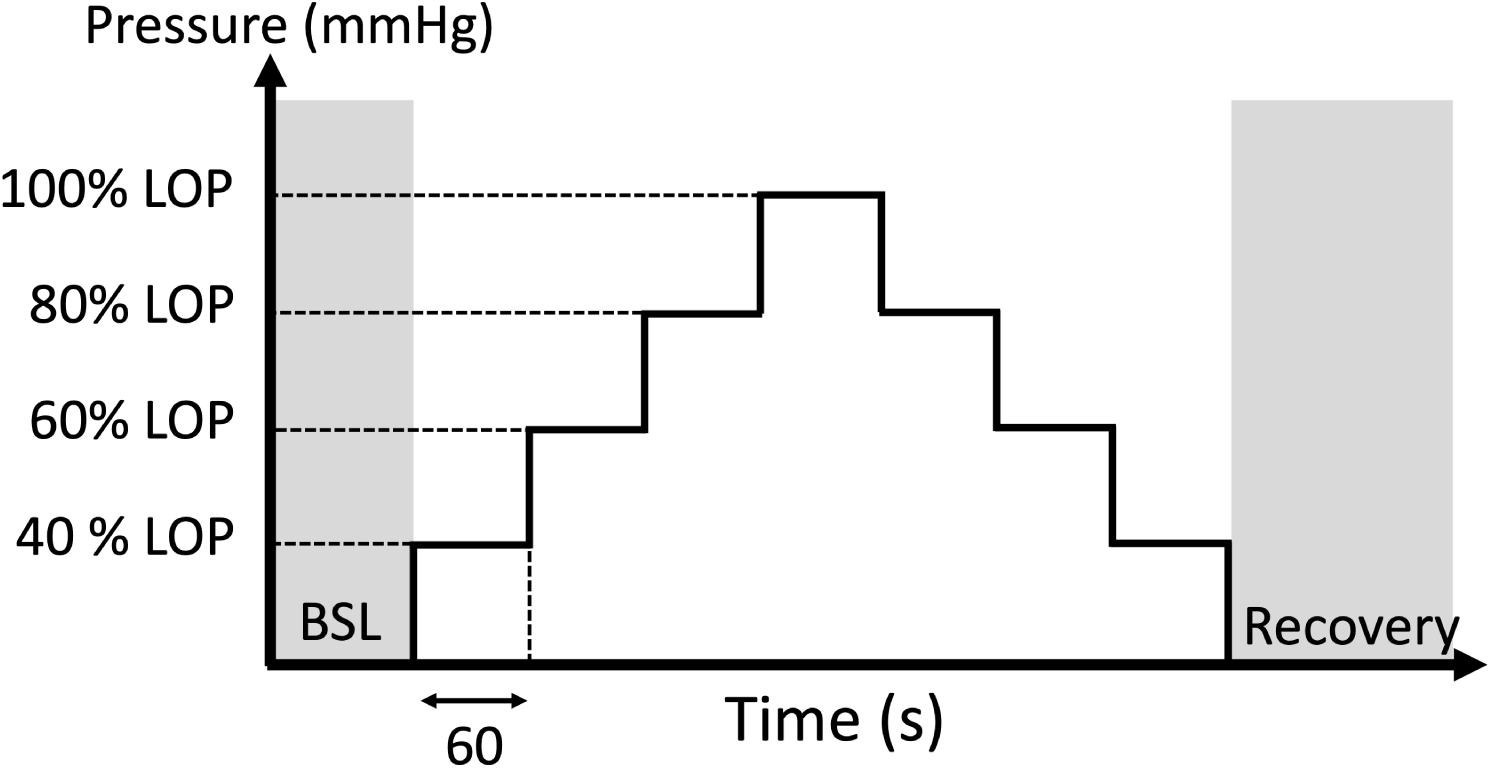
Schematic of the protocol for the assessment of the sensitivity of the platform *in vivo*. The cuff is inflated and deflated every 60 s at pressures equal to 40%, 60%, 80% and 100 % of the LOP. LOP: limb occlusion pressure; BSL: baseline.

#### 2.3.4 Optical interference

A specific ”quality phase” (QP) has been implemented to continuously assess the quality of TDNIRS, DCS, and pulse oximeter signals throughout the protocol. During this phase, the internal software alternates between switching DCS and TD-NIRS sources on and off to evaluate potential interference and dark light levels. This phase begins with signal equalization for TD-NIRS up to 10^6^ counts/s and adjusts DCS signals to the maximum deliverable power. The QP lasts less than two minutes, and its results are stored in data files and displayed to the user via quality indicators that turn red or green based on the outcome (Fig. 7).

**Fig 7.**
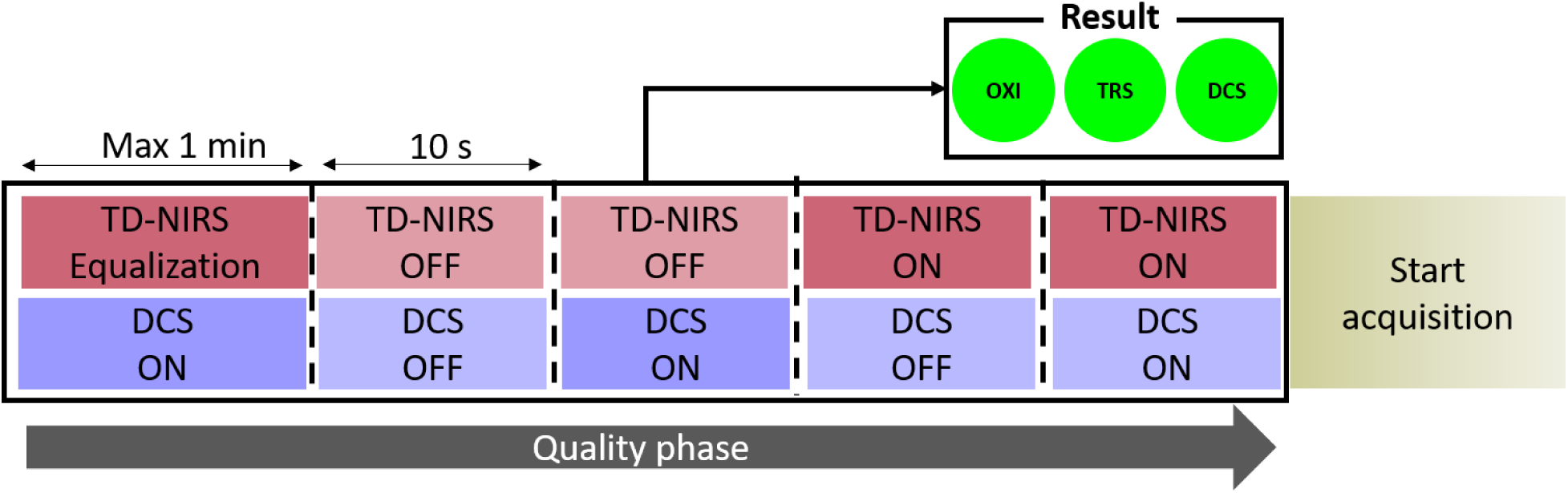
Schematic of data quality phase (QP). After an the inital equalization of the TD-NIRS, lasers are switched ON and OFF alternatively for a time interval of 10 s. The first minute correspond to the equalization phase of the TD-NIRS lasers.

At the end of the QP, the system flags any quality issues, such as low signal levels, interference between TD-NIRS and DCS, or poor pulse oximeter performance. These results guide operators in ensuring reliable data collection. Additional details about the software and its protocols have been discussed elsewhere.^86^

Approximately 10% of the subjects were randomly selected to investigate the influence of TDNIRS and DCS light levels on each other, both in the absence and presence of laser illumination. The background level was measured by integrating the total photon count when no diffuse light was incident on the detector, for both modules, both with and without the counterpart light active.

The effect of the counterpart light on the signal was assessed by integrating the photon count with the counterpart light either turned off or on. The influence of a module on the other was then analyzed using the Mann-Whitney test, with a significance level set at p*<*0.05.

### 2.4 Validation for clinical settings

#### 2.4.1 Comparison against commercially available device during a vascular occlusion test

The hDOS device was compared with a commercially available INVOS 5100C (Somanetic, Minnesota, USA) on two different muscles: the *palmaris longus* and *brachioradialis*. Subject demographics (age and biological sex) and the ATT were collected prior to the measurements. In particular, the ATT for both *palmaris longus* and *brachioradialis* muscles was measured by means of a portable ultrasound (ECUBEi7, ALPINION MEDICAL SYSTEMS Co., Ltd., Republic of Korea).

The INVOS 5100C and hDOS device were alternately positioned on one muscle or the other of healthy subjects in a randomized order for two subsequent VOTs, with an interval of approximately 30 minutes between each test.

For each measurement, the pertinent VOT-derived parameters, as explained previously, were calculated. In order to synchronize the two devices, manual markers were utilized that are then used as a reference in post processing.

The hDOS TD-NIRS module operates at 1 Hz, while the INVOS 5100C provides StO_2_ values every five to six s at its fastest rate. Therefore, TD-NIRS data were resampled to match the timing of the INVOS 5100C. The responses of the two devices to the VOT, as well as their performance across the two different muscles, were then compared.

The CV was used as a metric to assess intra-muscle variability within the two muscles. The statistical difference between the *palmaris longus* and *brachioradialis* was evaluated using the Wilcoxon signed-rank test, with significance set at p*<*0.05. Additionally, a Bland-Altman plot was employed to visualize and assess whether the StO_2_ measurements from the INVOS 5100C are interchangeable with those of hDOS device.

#### 2.4.2 Characterization of the microcirculation: comparison between healthy subjects and general mixed ICU patients

The clinical validation aimed to ensure the device’s efficacy and safety in critical care scenarios and it was part of the VASCOVID clinical campaign. Measurements on healthy subjects were conducted at the Institute of Photonic Sciences (ICFO) and were approved by the associated ethical committee (Ref: ICFO`_`HCP/2012/1).

The clinical protocol carried out at Parc Taulí Hospital received approval from its local Ethics Committee and the Comité d’Investigació amb Medicaments (CEIm) (Ref:2021/3015). The study was conducted in accordance with the Helsinki Declaration of 1975, and revised in 2008.

The purpose of this study is to characterize microvascular reactivity in the *brachioradialis* muscle of patients admitted to the ICU and compare it with healthy subjects.

The study includes two different groups: (1) Adult healthy volunteers with no previous history of disease that can affect blood circulation, recruited on a voluntary basis; (2) adult general ICU patients, including septic and non-septic patients.

Exclusion criteria for the ICU patients included the presence of venous thrombosis in the upper limbs, as well as hematomas or skin lesions on the forearm that could interfere with the placement of the hDOS device’s probe. Hemodynamically unstable patients, defined by uncorrected arterial hypotension or the need for active resuscitation to optimize blood pressure and/or cardiac output, were not included. Participation in the study was voluntary, with informed consent obtained either from the patient or from their legal representative.

Due to the exceptional situation of the pandemic from SARS-CoV-19 infection, if the physical access to the patients’ representatives was not possible, informed consent was obtained verbally, by means of telephone conference, with an external witness, not involved in the study. Data were collected and managed using REDCap (Research Electronic Data Capture) electronic data capture tools hosted at ICFO.^108, 109^

The vascular reactivity of the subjects was characterized by means of a VOT, as previously described in Section 2.2, on the *brachioradialis* muscle, with a pressure applied 50 mmHg above the systolic value. Systolic pressure was measured using a commercial blood pressure monitor on the contralateral arm for the healthy subjects, while in the ICU patients it was obtained by invasive blood pressure monitoring. Finally, the differences between the groups were evaluated by means of Mann-Whitney U test with a significance level of p*<*0.05.

## 3 Results

### 3.1 Precision of the hDOS device

#### 3.1.1 Variability of key parameters at module level

In a period of time of seven months, a total of 59 IRF measurements and corresponding phantom measurements were collected. These measurements correspond to 59 days of measurements corresponding to more than 100 sessions of measurements recorded at the hospital.

The CV for the FWHM in the IRF was 2.9% (1.1%) at 685 nm (828 nm), and for the barycenter, the precision was 3.9± 0.02 ns (CV = 0.4 %), and at 828 nm it was 3.6 ± 0.02 ns (CV = 0.6 %). The CV of the FWHM was 2.9 % for 685 nm and 1.1 % for 828 nm. For *µ*_a_, the CV was 0.9% (1.0%) at 685 nm (828 nm). For *µ*′_s_, the CV was 2.2% (2.3%) at 685 nm (828 nm). When calculating the effective tHb^ph^, the CV was 1.0%, and for StO_2_^ph^, it was 1.2% over the entire measurement period. The intra-phantom variability was 1.4% for tHb^ph^ and 1.3% for StO_2_. Results for the effective tHb^ph^ and StO_2_^ph^ are shown in the Fig. 8 where they have been grouped by month for clarity and a summary of the results is reported in the Table 1. Each dot represents the mean and the errorbars the standard deviation over 20 consecutive repetitions. The shaded areas represent the standard deviation over the whole period of time considered.

**Table 1.** Coefficient of variation (CV %) calculated across all measurement sessions recorded in seven months for both the full-width-half-maximum (FWHM) of the IRF and the optical properties phantom (*µ*_a_ in cm*^−^*^1^ and *µ′*_s_ in cm*^−^*^1^). For the barycenter the standard deviation across all measurement sessions is reported, in ns. A total of 59 IRF and phantom measurements were collected during this period. The *µ*_a_ in cm*^−^*^1^ was employed to calculate the effective tHb^ph^ and StO_2_^ph^.

| IRF |  |  | Phantom |  |  |  |
| --- | --- | --- | --- | --- | --- | --- |
| | FWHM (%) | Barycenter (ns) | $\mu_a$ (%) | $\mu'_s$ (%) | $\text{tHb}^{\text{ph}}$ (%) | $\text{StO}_2^{\text{ph}}$ (%) |
| 685 nm | 2.9 | 0.02 | 0.9 | 2.2 | 1.0 | 1.2 |
| 828 nm | 1.1 | 0.02 | 1.0 | 2.3 |  |  |

**Fig 8.**
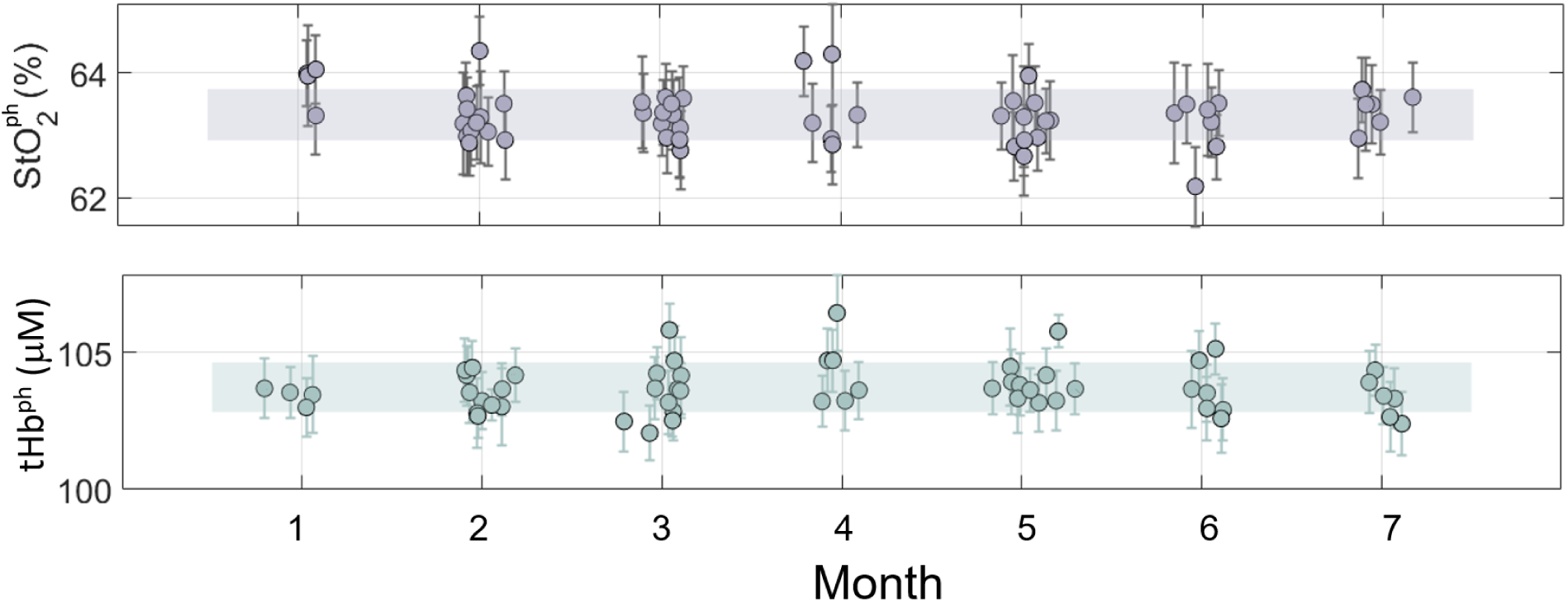
Effective StO_2_^ph^ in %(top panel) and tHb^ph^ in *µ*M (bottom panel) obtained by the solid phantom for 59 days of measurements, grouped by month for clarity. The dots represent the average values while the error bars represent the standard deviation over 20 repetitions. The shaded areas represent the standard deviation over all the measurements.

**Fig 9.**
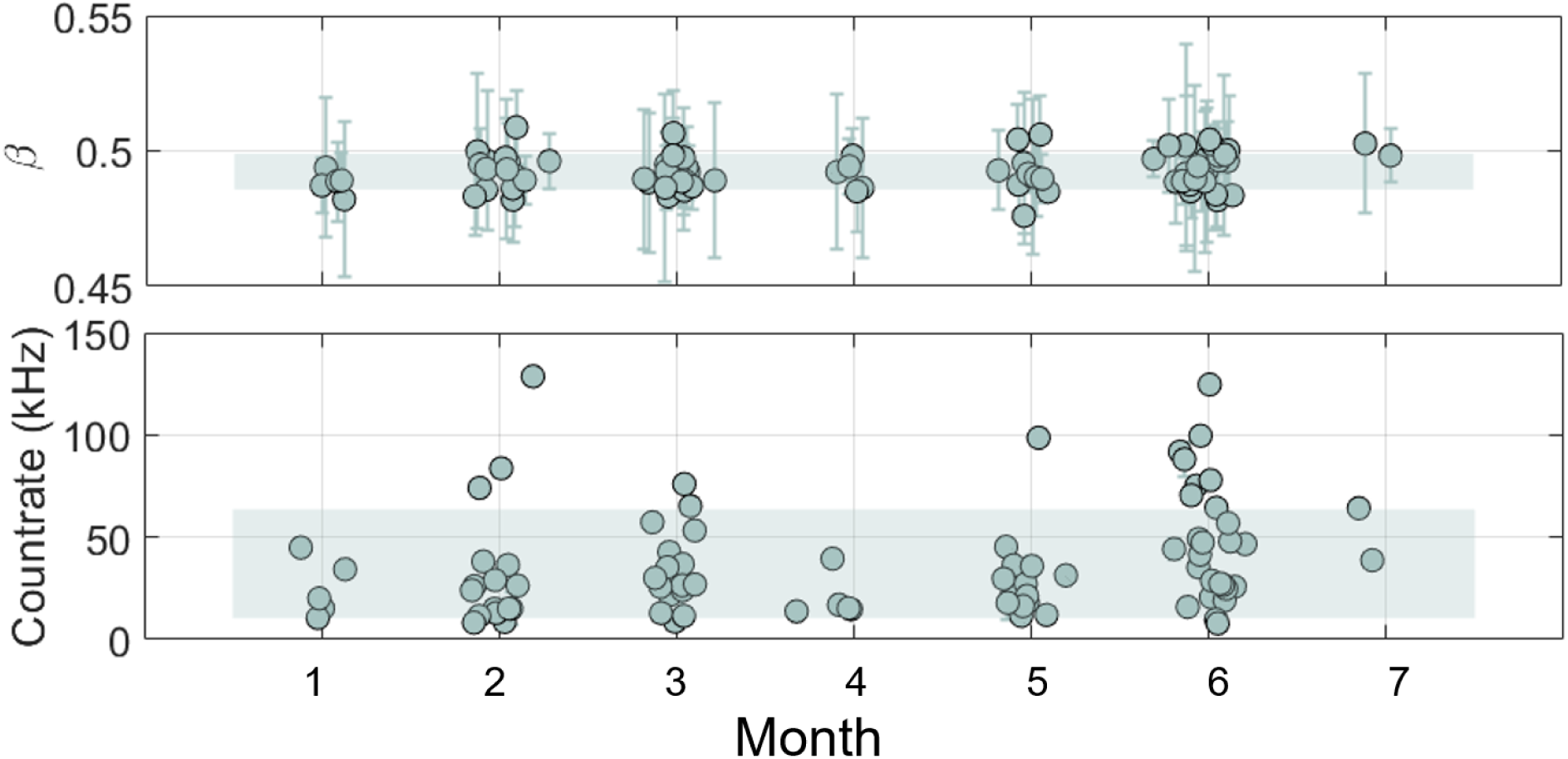
Top panel: *β* parameter obtained for seven months worth of measurement. The dots represent the average values over 60 s of baseline, with averaging time of 10 s, while the error bars represent the standard deviation. Botton panel: count rate for DCS in kHz for each subject admitted in the clinical campaign. Dots represent the mean values, while errorbars the standard deviation over 20 s of baseline measurement. The green shaded area correspons to the standard deviation over all the measurements.

When fitting each phantom with the first phantom of the month (e.g., all phantom measurements acquired in month 3 are fitted with the first IRF acquired in month 3), we obtained a higher, but not statistically significantly different, variability in the retrieval of *µ*_a_ (CV = 2.8% for both 685 and 828 nm; p = 0.52) and *µ*′_s_ (CV = 2.5% for both 685 and 828 nm; p = 0.75). This higher variability translated to a CV = 2.8% in the effective tHb^ph^ and a CV = 1.0% in the StO_2_^ph^. More details about the variability on the optical properties of the phantom are shown in Appendix 6.1.

The photon count rate and *β* parameter were analyzed for N = 137 subjects. The photon count rate was subject-dependent and showed a weak positive trend (Spearman’s correlation, *ρ* = 0.3, p = 0.03). Since *in vivo* conditions vary across subjects, the recruitement of subjects with better signal might have contributed to this weak positive trend. The *β* parameter had an average value of *β* = 0.49 ± 0.01 with a CV = 1.9%, recorded in seven months worth of measurements. No significant trends were observed for *β* over time (Spearman’s correlation, *ρ* = 0.2, p = 0.06). This result suggests that there are no degradation in the system over the time of measurements at the hospital.

Further details about DCS quality parameters are available in Appendix 6.1.

#### 3.1.2 Test-retest variability

As a test-retest evaluation, the hDOS device’s probe was replaced five times onto the *brachioradialis* muscle of a healthy female subject (32 years old). The results are summarized in Fig. 10. The mean values of StO_2_ and BFI across the five tests are reported. An inter-repetition CV of 1.2% and CV of 12.6% were obtained for the entire experiment. The within-test variability was found to be CV *<* 0.8% for StO_2_ and CV *<* 15.2% for BFI and it was similar to what was found in Ref.,^103^ although StO_2_ was better with hDOS (1.2% reported here compared to 8.5% in Ref.^103^).

**Fig 10.**
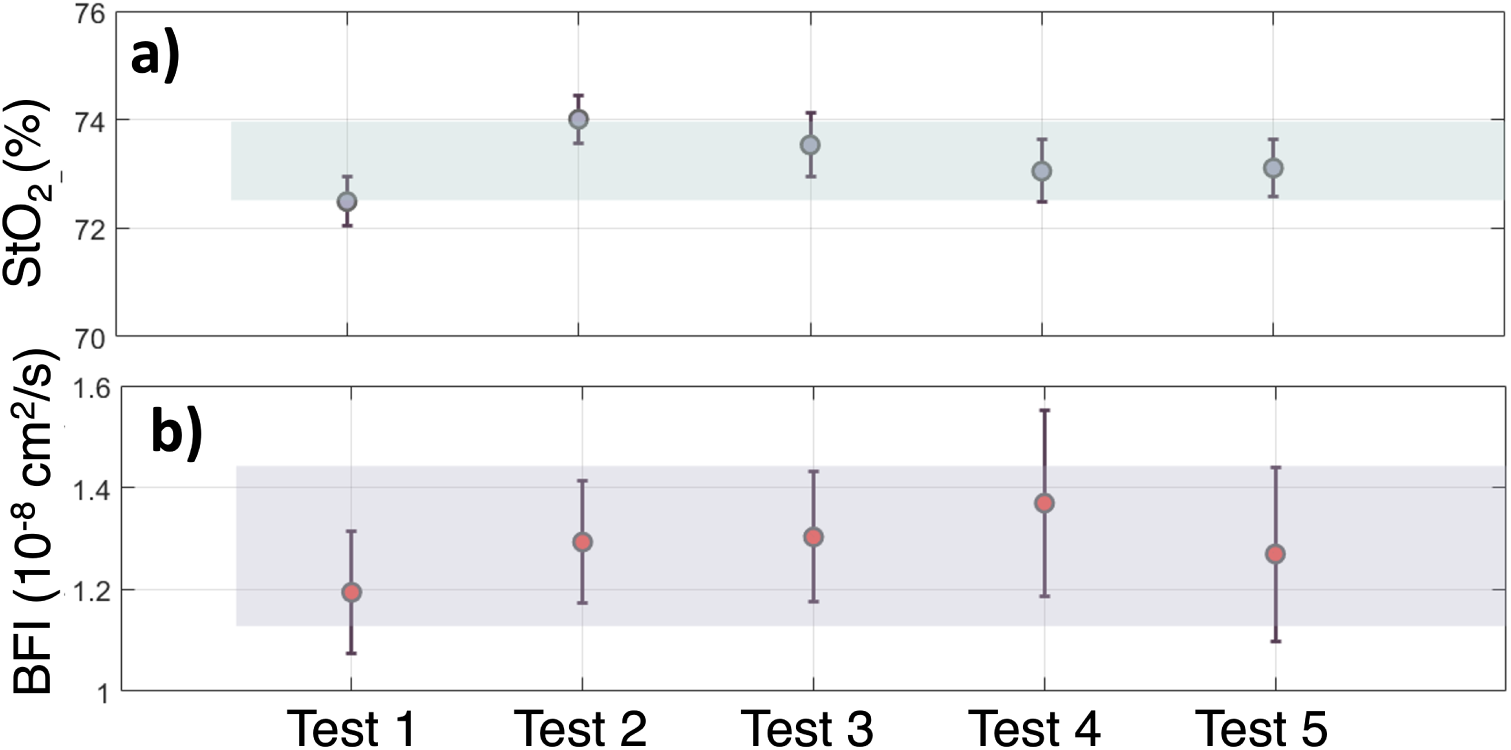
a) StO_2_ values obtained over 5 repositioning. The dots represent the mean values, while the errorbars are the standard deviations calculated over 200 acquisitions. The shaded area represent the standard deviation, centered around the mean over the 5 tests.

#### 3.1.3 Dependence of precision with respect to absolute values

Seven (N=7) healthy subjects were enrolled to evaluate the variability at different light levels of both StO_2_ and BFI. In Fig. 11 an example is reported for a single subject. As displayed in Fig. 12, it can be observed that for both BFI and StO_2_, the CV was dependent on the mean value retrieved during the various periods of inflation/deflation, with higher CV observed during deflation for both StO_2_ and BFI (panels a and c, respectively). The CV BFI exhibited similar behavior, showing smaller variability around higher count-rates (panel b). As expected, CV StO_2_ decreased with higher dynamic ranges of the acquired TD-NIRS curves (panel d). A statistically significant correlation was obtained between the CV BFI and count rate (*ρ* = -0.41, p = 0.001). No statistically significant correlation was found between count rate and the increase in BFI (*ρ* = -0.09, p = 0.46), nor between CV BFI and BFI (*ρ* = -0.19, p = 0.15). For StO_2_, a statistically significant correlation was found between CV StO_2_ and the dynamic range (DR) at 685 nm (*ρ* = -0.5, p *<* 0.001), as well as with StO_2_ (*ρ* = -0.33, p = 0.007). Finally, no statistically significant correlation was found when comparing StO_2_ and the dynamic range at 685 nm (*ρ* = 0.08, p = 0.5).

**Fig 11.**
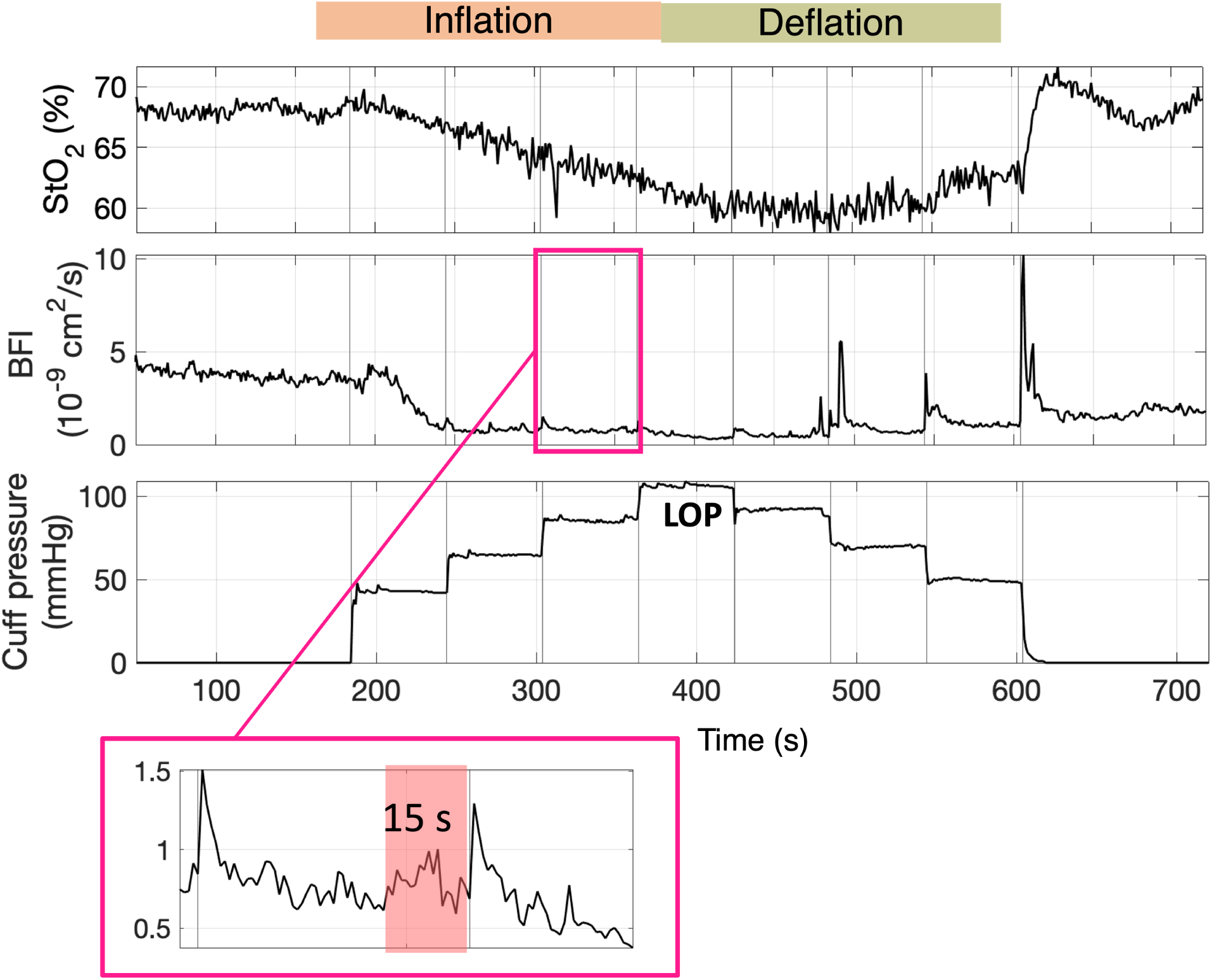
Example of VOT-LOP protocol for tissue oxygen saturation (StO_2_ in % and bloof flow index (BFI in cm^2^/s). The cuff was inflated and deflated in step of 40,60,80 and 100 % of the LOP, with occluding pressure maintained for 1 minute each time. Mean value, standard deviation and CV are calculated in the last 15 s of occlusion for each interval.

**Fig 12.**
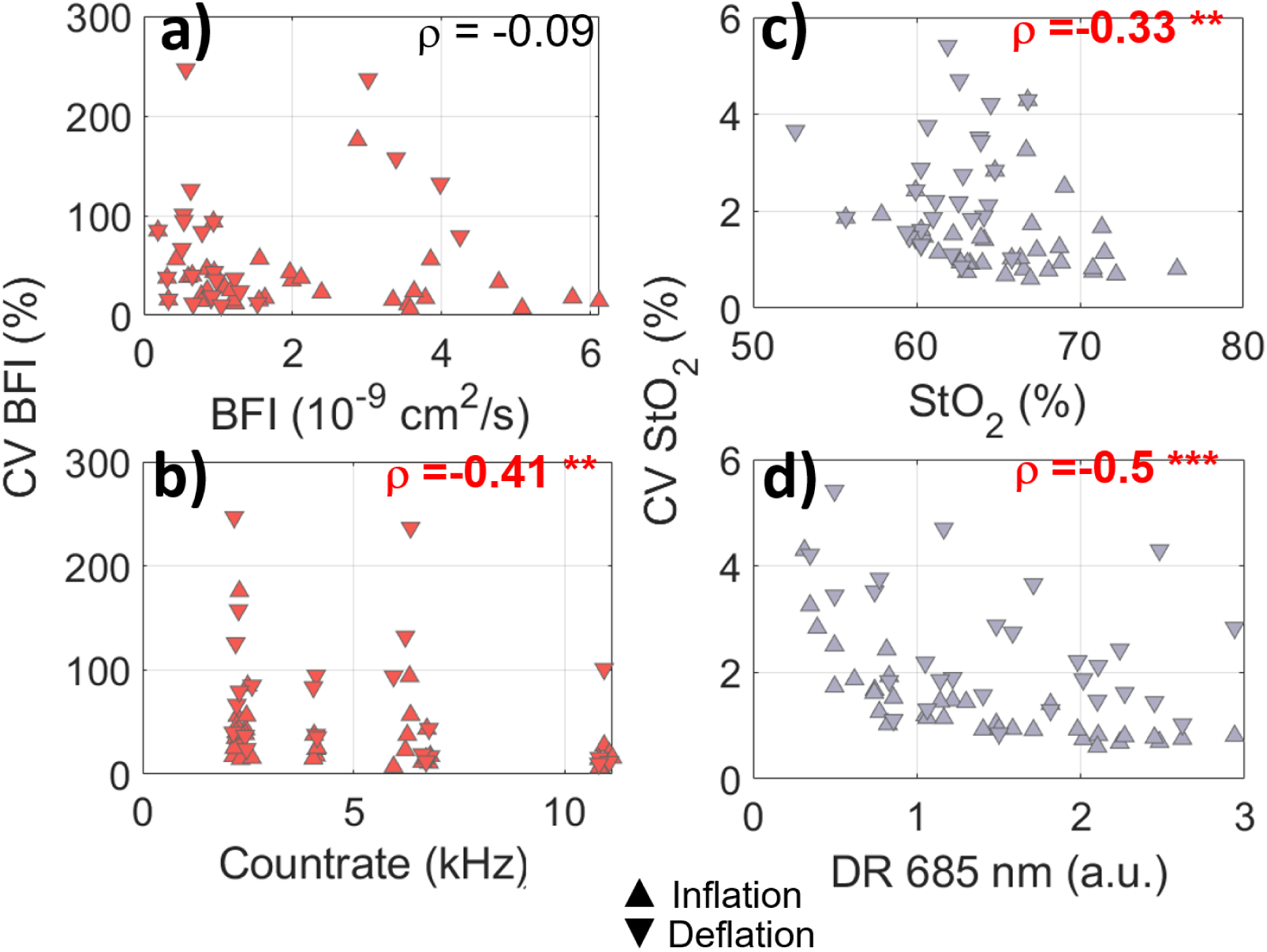
Coefficient of variation (CV in %) of blood flow (BFI) with respect to a) the mean values of BFI (in cm^2^/s) and b) to the count rate (in kHz), retrieved during the inflation/deflation periods; CV of tissue oxygen saturation (StO_2_) with respect to c) the mean value of StO_2_ (in %) and d) the dynamic range (DR) of the wavelength 685 nm.

#### 3.1.4 Optical interference

Results for the assessment of the presence of optical interference, is reported in Fig. 13.

**Fig 13.**
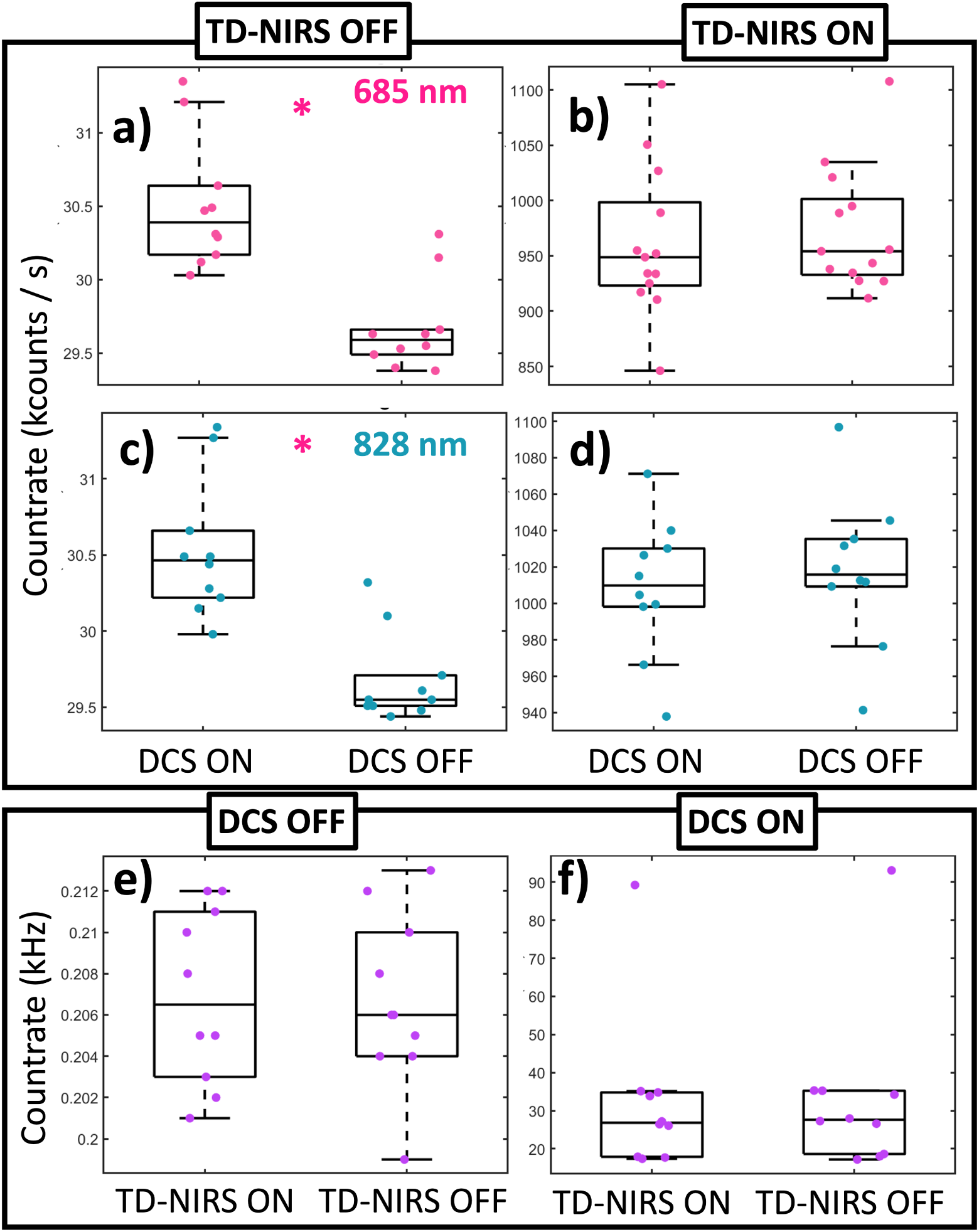
Boxplots illustrating the influence of the DCS signal on the TD-NIRS total background count rate (TD-NIRS OFF) when the DCS was ON or OFF for wavelength 685 (panel a) and 828 (panel c) nm; b) influence of the DCS signal on the total count rate of the TD-NIRS module (TD-NIRS ON) when the DCS was ON or OFF for the wavelengths 685 (panel b) and 828 (panel d). Influence of the TD-NIRS signal when the TD-NIRS lasers are either ON or OFF, when the DCS lasers was either OFF (panel e) or ON (panel f). A statistically significant difference is depicted by “*” when p*<*0.05.

The background count rate for the TD-NIRS was significantly higher when the DCS laser was emitting at full power (p*<*0.001 for both 685 and 828 nm). On the other hand, the background count rate of the DCS is not affected by the TD-NIRS lasers shining (p=0.48). Finally, a difference in the count rate when the TD-NIRS lasers are emitting and the DCS was either ON or OFF has not been found (p=0.15 for 685 nm and p=0.60 for 828 nm). Similar results are obtained when the DCS laser is emitting, and the TD-NIRS lasers are switched ON and OFF (p=0.31). The results suggest that the filters are functioning as expected, effectively preventing significant interference. The increase in background count rate when the DCS was ON, though statistically significant, was minimal and spread across the entire temporal window. Despite this increase, the system’s performance remains uncompromised, as sufficient dynamic range was maintained to reliably fit the DTOF and extract robust optical properties from the signal. A representative result is shown in Figure 14 where the StO_2_ and the BFI of a single patient is reported during the automatic QP. In this particular case, the CV for the BFI was 7.5% when the DCS laser was ON while the TD-NIRS lasers were OFF. Conversely, when the TD-NIRS module was ON and the DCS was OFF, the CV for the *µ*_a_ and *µ*′_s_ was less than 1% at both wavelengths, with a resulting CV for StO_2_ at 0.6% and for tHb at 0.5%. When both the TD-NIRS and DCS modules were ON, the CV for BFI remained at 7.5%, while the CV for StO_2_ and tHb increased slightly to 0.8%, and the CV for *µ*_a_ and *µ*′_s_ was less than 1.2% at both wavelengths.

**Fig 14.**
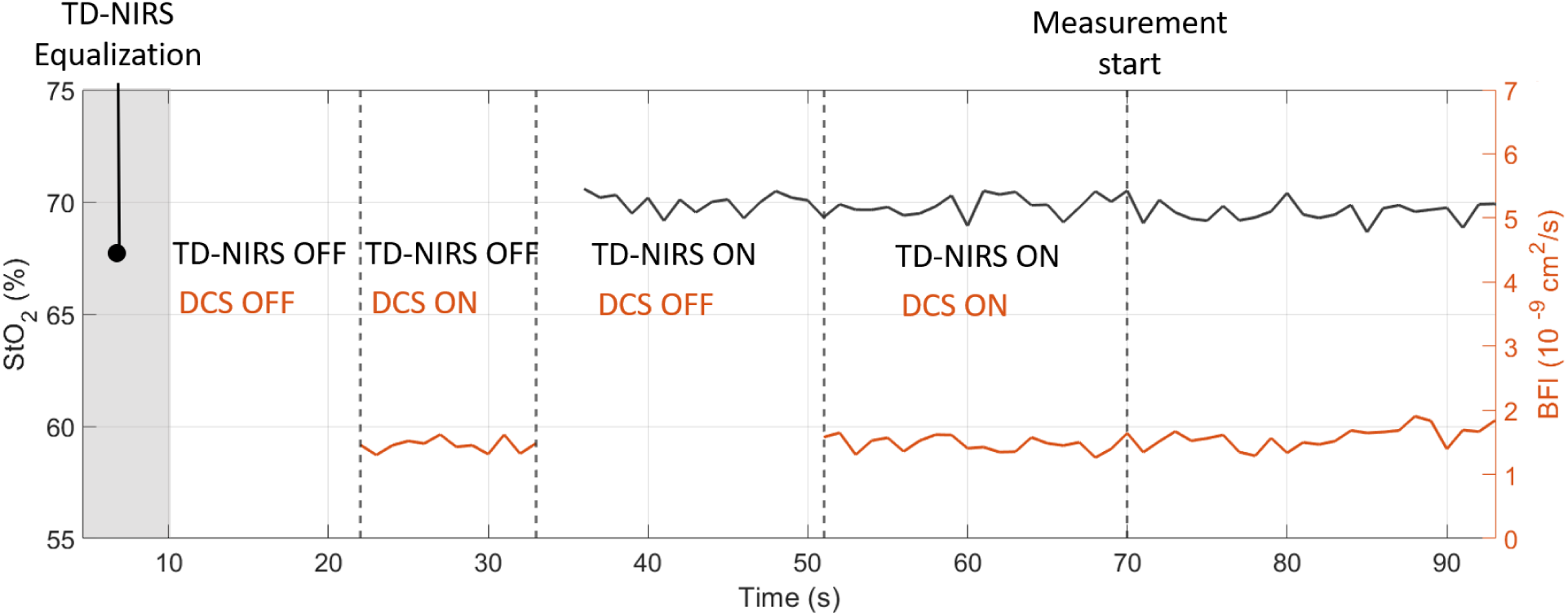
Example of StO_2_ (in black solid line) and BFI (in orange solid line) time traces during the quality phase. Dashed vertical llines denote the changes in ON/OFF cycles. A grey shaded area represents the equalization phase for TD-NIRS to reach the desired count rate 10^6^ counts/s.

### 3.2 Validation for clinical settings

#### 3.2.1 Comparison against a commercially available device

A total of ten subjects (six females) with a mean age of 28±5 years old and a BMI of 24.9±2.3 kg/m^2^ were included in this study. In addition, the ATT over the two measured muscles was 4.0±0.1 mm for the *brachioradialis* and 4.4±0.1 mm for the *palmaris longus* which were not statistically significantly different from each other (p=0.67). An example of hDOS device and INVOS 5100C time traces is reported in the Fig 15. The response to the ischemic challenge was lower in the hDOS device. Moreover, the StO_2_ in the INVOS 5100C was limited to a maximum value of 95% and a minimum value of 15%, which makes the extraction of the area under the curve unreliable (see the zoomed area in Fig 15). A summary of the VOT-derived parameters, their medians and interquartile ranges are shown in Table 2 for both devices and muscles.

**Fig 15.**
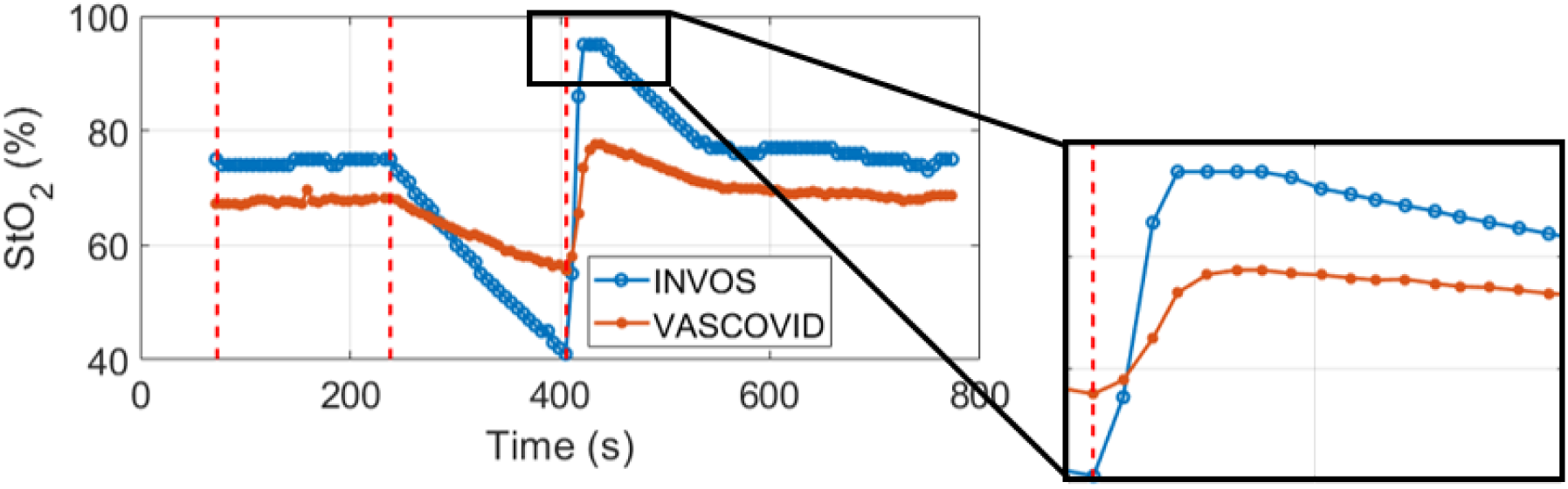
Example of the time trace of a simultaneous measurement of hDOS device (orange line resampled at the INVOS 5100C sampling time) and INVOS 5100C (blue line). The dashed lines represent the start of the measurement, after the data quality phase, inflation and deflation time. The StO_2_ hyperemic peak is highlighted in the zoomed window.

**Table 2.** Comparison of hDOS TD-NIRS and INVOS 5100C values obtained for the *brachioradialis* and *palmaris longus* during the VOT. Values are median and first and third interquartile range values in square brackets. The symbol “*” represents statistically signficant difference between *brachioradialis* and *palmaris longus*; The symbol “†” represents statistically significant difference between the hDOS TD-NIRS and INVOS 5100C. In both cases the significance was set for p*<*0.05.

|  | hDOS TD-NIRS |  | INVOS 5100C |  |
| --- | --- | --- | --- | --- |
|  | Brachioradialis | Palmaris longus | Brachioradialis | Palmaris longus |
| StO<sub>2</sub> (%) | 72.0 [69.9-72.5]<sup>†</sup> | 70.7 [68.4-73.0] | 76.4 [72.8-79.4] | 74.0 [69.0-81.7] |
| DeO<sub>2</sub> (%/min) | -4.6 [-5.2- -3.7] *,<sup>†</sup> | -5.4 [-6.6- -4.4]<sup>†</sup> | -10.9 [-14.4- -8.9] | -11.4 [-13.6- -10.3] |
| ReO<sub>2</sub> (%/s) | 0.9 [0.7-1.2] *,<sup>†</sup> | 1.2 [1.0-1.6] <sup>†</sup> | 2.7 [2.2-3.7] | 2.8 [2.3-3.1] |
| AUC StO<sub>2</sub> (%· min) | 6.4 [3.6-8.5] | 7.4 [5.6-7.7] | 7.5 [4.9-8.5] | 7.4 [6.6-8.8] |

The hDOS TD-NIRS StO_2_ presents a smaller interquartile range, particularly in DeO_2_ and ReO_2_. Concerning intersubject variability, a CV of 3.3 (5.0)% for hDOS device in the *brachioradialis* (*palmaris longus*) was obtained, compared to 6.1 (9.6)% for INVOS 5100C. Additionally, when intrasubject variability was considered, an average of 0.4 (0.6)% was found for hDOS device versus 0.8 (1.0)% for the *brachioradialis* (*palmaris longus*) in INVOS 5100C, respectively. Statistical differences in the positioning of the probe were not observed for any of the variables retrieved by INVOS 5100C. However, differences were found in the DeO_2_ (p = 0.004) and ReO_2_ (p = 0.009) variables between the *brachioradialis* and *palmaris longus* when examining TD-NIRS-hDOS. In the *brachioradialis*, differences were noted in the StO_2_ baseline (p = 0.02), as well as in DeO_2_ (p*<* 0.001) and ReO_2_ (p *<* 0.001) when comparing TD-NIRS and INVOS 5100C. No significant differences were found in the AUC StO_2_ (p = 0.61). In the *palmaris longus*, differences were observed only in DeO_2_ and ReO_2_ (p *<* 0.001).

In Fig. 16 a Bland-Altman plot is reported where for each subject (in different colors), the differences between INVOS 5100C and hDOS StO_2_ are plotted against their average values at each time point of the VOT. The black solid line correspond to the bias, while the black dashed lines correspond to ±1.96 times the standard deviation. A bias of +2.14% is reported which is not representative in this case since there is a non-zero slope (R = 0.72, p*<*0.001) confirming that the two devices differ from each other in a non-trivial manner. Furthermore, this difference is not subject dependent. In fact, all subjects display a significant non-zero slope.

**Fig 16.**
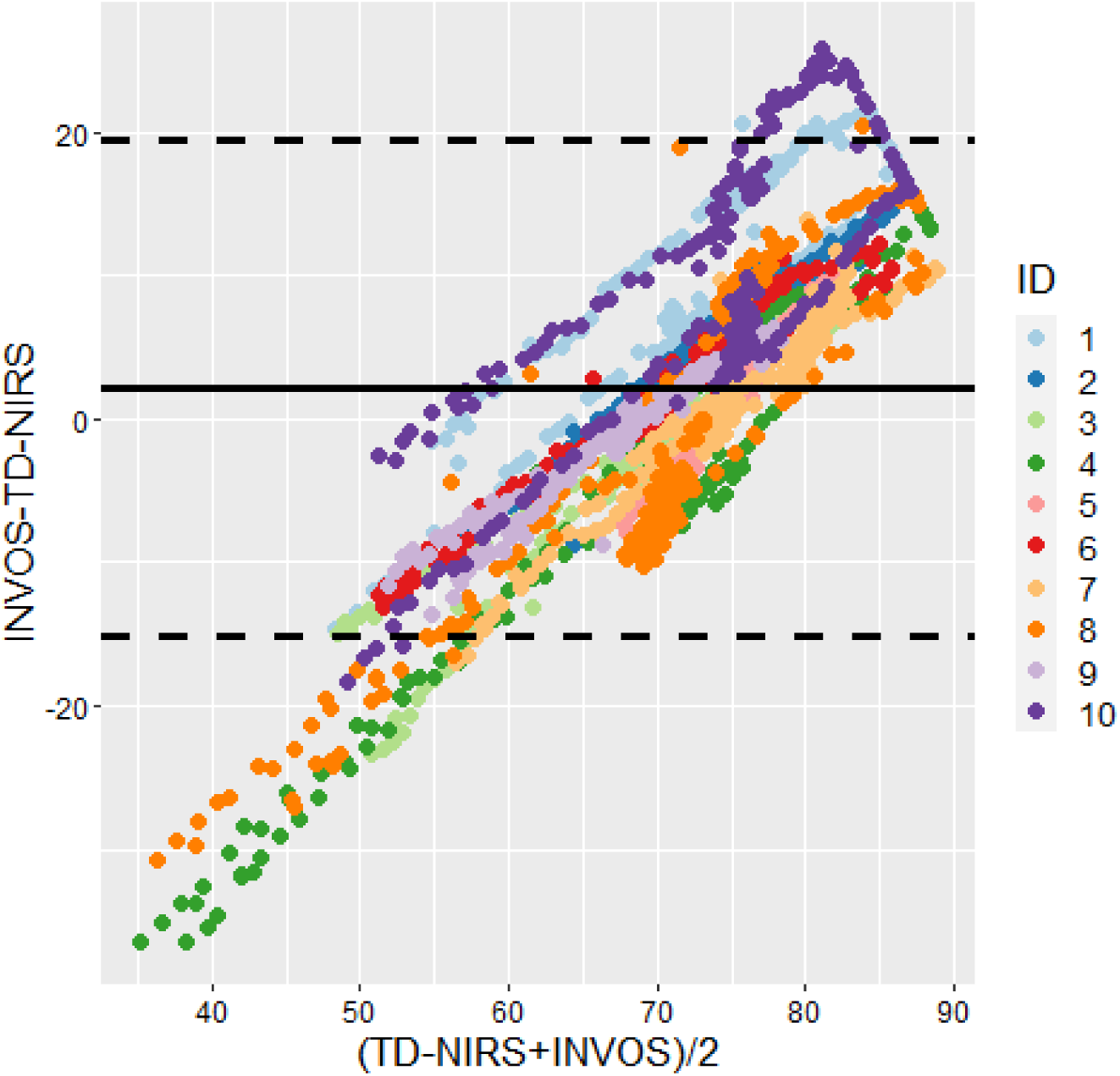
Bland-Altman plot for StO_2_ as measured by INVOS 5100C (indicated as INVOS) and hDOS TD-NIRS (indicated as TD-NIRS). Colors represent all the subjects ID included in the study (N=10). Both positions of measurement (*brachioradialis* and *palmaris longus*) are taken into consideration in the plot.

#### 3.2.2 Characterization of the microcirculation: comparison between healthy subjects and general mixed ICU patients

Thirty-seven (N=37) healthy subjects and one hundred (N=100) general ICU patients were recruited. The demographic, clinical, and morphological data are summarized in Table 3, while the optical data, as well as baseline and median responses to the ischemic challenge, are presented in Table 4.

**Table 3.**
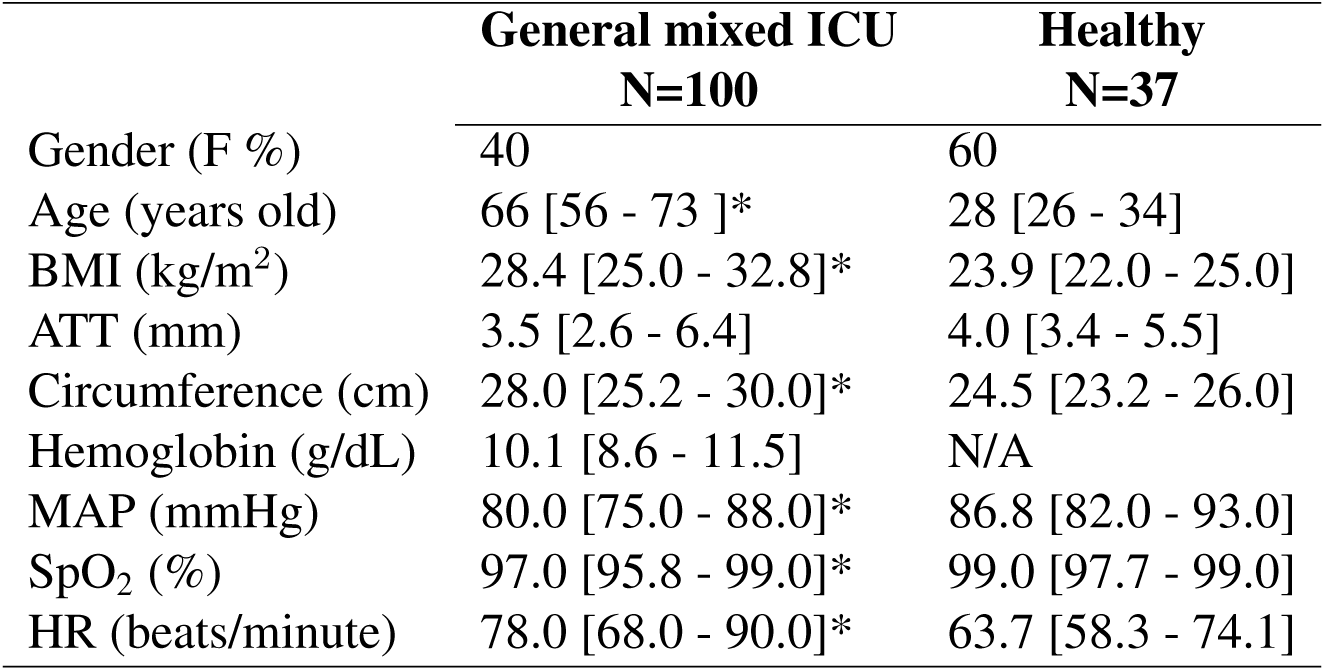
Demographic data. Median and interquartile range (25% and 75%) is reported in square brackets. BMI: body mass index; ATT: adipose tissue thickness; MAP: mean arterial pressure; SpO_2_: peripheral tissue oxygenation; HR: heart rate; N/A: information not available or not applicable. (*) indicates difference between the general mixed ICU and healthy population with a significance level p*<* 0.05 according to Wilcoxon rank-sum test.

|  | **General mixed ICU**<br>**N=100** | **Healthy**<br>**N=37** |
| --- | --- | --- |
| Gender (F %) | 40 | 60 |
| Age (years old) | 66 [56 - 73 ]* | 28 [26 - 34] |
| BMI (kg/m<sup>2</sup>) | 28.4 [25.0 - 32.8]* | 23.9 [22.0 - 25.0] |
| ATT (mm) | 3.5 [2.6 - 6.4] | 4.0 [3.4 - 5.5] |
| Circumference (cm) | 28.0 [25.2 - 30.0]* | 24.5 [23.2 - 26.0] |
| Hemoglobin (g/dL) | 10.1 [8.6 - 11.5] | N/A |
| MAP (mmHg) | 80.0 [75.0 - 88.0]* | 86.8 [82.0 - 93.0] |
| SpO<sub>2</sub> (%) | 97.0 [95.8 - 99.0]* | 99.0 [97.7 - 99.0] |
| HR (beats/minute) | 78.0 [68.0 - 90.0]* | 63.7 [58.3 - 74.1] |

**Table 4.** Optical data obtained for the three subjects’ cohorts. The mean values and standard deviations (mean ± std. dev) of tHb, StO_2_, BFI, OEF and MMRO_2_ are calculated over 30 s prior to the occlusion interval. Median and interquartile range (25% and 75%) of DeO_2_, ReO_2_, AUC StO_2_ and AUC BFI are reported in square brackets. (*) indicates difference between the general mixed ICU and healthy population with a significance level p*<* 0.05 according to Wilcoxon rank-sum test.

|  | **General Mixed ICU**<br>**N=100** | **Healthy**<br>**N=37** |
| --- | --- | --- |
| $\mu_a$ 685 (cm<sup>-1</sup>) | 0.22 ± 0.07 | 0.22 ± 0.06 |
| $\mu_a$ 828 (cm<sup>-1</sup>) | 0.21 ± 0.05 | 0.23 ± 0.06 |
| $\mu'_s$ 685 (cm<sup>-1</sup>) | 9.25 ± 1.11 * | 8.62 ± 0.77 |
| $\mu'_s$ 828 (cm<sup>-1</sup>) | 8.65 ± 1.22 * | 8.18 ± 0.85 |
| tHb (μM) | 104.04 ± 26.75 | 112.88 ± 27.82 |
| StO<sub>2</sub> (%) | 67.05 ± 5.54 * | 70.33 ± 3.73 |
| BFI (10<sup>-8</sup> cm<sup>2</sup>/s) | 0.49 ± 0.50 * | 0.53 ± 0.31 |
| OEF (%) | 30 ± 6* | 28 ± 4 |
| MMRO<sub>2</sub> (10<sup>-8</sup> % · cm<sup>2</sup>/s) | 1.39 ± 1.23 * | 2.09 ± 1.1 |
| DeO<sub>2</sub> (%/min) | -4.3 [-5.7- -3.3] | -4.1 [-5.2 - -3.1] |
| ReO<sub>2</sub> (%/s) | 0.8 [0.5- 1.1] * | 1.1 [0.9 - 1.6] |
| AUC StO<sub>2</sub> (·min) | 3.8 [2.0 - 6.5] * | 7.7 [5.2 - 9.3] |
| AUC BFI (10<sup>-6</sup>cm<sup>2</sup>) | 2.2 [1.1 - 3.3] * | 4.0 [2.6 - 5.7] |

It has been found statistically significant difference between the general ICU and healthy groups in age, BMI, arm circumference, as well as SpO_2_ and HR at baseline (p*<* 0.001). MAP also showed a signficant difference between the two groups (p=0.02).

Significant differences were found in *µ*′_s_ at both wavelengths (p*<*0.001). Healthy group showed a significantly higher StO_2_ with respect to the general mixed ICU group (p*<*0.001). A significant difference in BFI was detected (p=0.03) with higher values in the healthy group. MMRO_2_ was found to be significantly reduced in the ICU population (p*<*0.001). During the VOT, significantly lower values were identified for the ReO_2_, AUC StO_2_ and AUC BFI in the ICU population compared to healthy subjects (all p*<*0.001).

## 4 Discussion

In this paper, the hDOS device is introduced as a versatile, multimodal system that integrates advanced TD-NIRS and DCS technologies, along with essential accessories such as a pulse oximeter and an automated tourniquet for vascular occlusion tests and baseline metabolism assessment.

The validation of this platform went beyond basic performance metrics, emphasizing its robustness and suitability for independent clinical use. While optimized for intensive care, its adaptability extends to various settings, including laboratory environments, clinics, and more demanding conditions such as the operating room and emergency care unit.

The device underwent rigorous testing, with over 200 hours of use across approximately 150 measurement sessions.

A user-friendly software improves usability with an intuitive interface, quality feedback, and real-time safety checks to ensure compliance with safety standards. The multimodal probe, equipped with a force sensor, touch sensor, and accelerometer, aids in standardizing probe pressure, improving measurement quality, and enabling data rejection when necessary. The accelerometer detects motion artifacts, while the capacitive touch sensor ensures continuous tissue contact for accurate measurements.^89, 110, 111^

Deployed in the intensive care unit for seven months ^2^, the device underwent evaluations including tests on tissue-mimicking phantoms and *in vivo* assessments to gauge its stability, reproducibility, and performance in a clinical setting. To note that for some of these assessments a replica device has been utilized. These replicas differ only in having slightly improved and more robust electronics, but are optically identical. Specifically, it has been the focus of the work of Ref.^12^ where a comprehensive comparison involving ten TD-NIRS devices based on the same hDOS TD-NIRS technology was presented. They in fact revealed a remarkably high level of reproducibility and accuracy in the retrieval of optical parameters from well-characterized tissue-mimicking phantoms among different replicas, which is promising for the consistency and reliability of TD-NIRS devices manufactured at scale and utilized in this very same platform. In particular, for a similar system, in Ref.^112^ authors report a variability better than 1.2% on different types of phantoms and different, bulkier implementations of TD-NIRS technology, with comparable optical properties to the one employed with hDOS device.

For hDOS TD-NIRS a variability in tHb^ph^ and StO_2_^ph^ was found to be below 1.2%. The focus was on assessing a day-to-day variability, not on validating absolute values. These results are consistent with findings from Ref.,^103^ where variability was 3.0% over nearly 10 months. Monthly calibration with the IRF showed less than 3% variability in effective tHb^ph^ and StO_2_^ph^, despite minor, statistically insignificant variations in optical properties (*µ*_a_ and *µ*′_s_).

DCS stability, well-documented in Refs.,^18, 20, 21, 88, 91, 113–115^ was confirmed by monitoring count rate and *β* parameters in *in vivo* measurements. No significant trends were observed, indicating stable performance during data acquisition.

The precision of the hDOS device was evaluated through test-retest and precision analysis concerning absolute values. In a test-retest study on a single subject, TD-NIRS results were consistent with literature for both TD-NIRS^95, 116^ and DCS.^87, 117^ The CV for StO_2_ with hDOS was notably lower at 1.2% compared to 8.5 % reported previously^103^ and also superior to MOXY (Fortiori Design LLC, Minnesota, US) and Portamon (PortaMon, Artinis, Medical System, The Netherlands) devices, which reported CVs of less than 2.5%.^118^ The hDOS TD-NIRS shows a better precision with respect to a novel commercially available wearable device (Train.Red FYER) where a CV of 5% was reported.

Regarding the evaluation of the precision with respect to absolute values, CV was found to be dependent on light levels for both StO_2_ and BFI. During dynamic phases, such as deoxygenation and reoxygenation, a lower StO_2_ resulted in a reduced SNR and increased CV. Future work will aim to improve precision across all measurement ranges using real-time optimization algorithms and fast tunable filters. Despite this, the precision was better than 4% across the StO_2_ range. BFI variability increased during full arterial occlusion, but this does not impact measurements as BFI during occlusion only confirms ischemia. Evaluating *in vivo* variability, especially in skeletal muscle, remains complex and underexplored compared to brain studies.

Precision studies in tissue mimicking phantoms and *in vivo* provide a valuable benchmark when translating the usefulness of these technologies to clinical applications. On the other hand, it is also critical to assess how it compares to a less expensive commercially available device, such as the the INVOS 5100C. The INVOS 5100C, like the hDOS TD-NIRS system, uses near-infrared light for tissue oxygenation assessment but relies on the so-called SRS algorithm/probe.^1, 2^ While valued for its cost-effectiveness and ease of use, it only measures tissue oxygenation without providing direct indicators of perfusion and metabolism. Variability in findings from NIRS-VOT studies^119^ underscores the challenge of uniform standards, an issue addressed in the *in vivo* validation of the VASCOVID project. The key difference between these technologies lies in their assumptions: the INVOS 5100C assumes light attenuation is primarily due to absorption and that scattering depends linearly on wavelength,^120, 121^ deriving a tissue saturation index. In contrast, the TDNIRS system in the hDOS device calculates absolute absorption values without such assumptions, overcoming limitations of light penetration depth by adjusting injected power within safety limits. Literature consensus supports that TD-NIRS offers superior accuracy, precision, and repeatability compared to CW-NIRS methods.^97, 122^ This suggests that clinical systems like the INVOS 5100C may provide less accurate data, especially during vascular occlusion tests. While definitive *in vivo* comparisons are challenging due to varying algorithms and lack of gold standards, hDOS TD-NIRS demonstrated greater consistency with lower intersubject and intrasubject variability when comparing baseline StO_2_ to INVOS 5100C. Statistical differences were observed in VOT-derived parameters when comparing the *brachioradialis* and *palmaris longus* positions for INVOS 5100C but not for hDOS TD-NIRS. hDOS TD-NIRS showed distinctions in DeO_2_ and ReO_2_ between probe positions. Specifically, in the *brachioradialis*, differences in StO_2_ baseline, DeO_2_, and ReO_2_ were noted, while in the *palmaris longus*, differences were seen in DeO_2_ and ReO_2_. A Bland-Altman plot analysis revealed a 2.1% which was not representative due to a non-zero slope. This highlights that the two devices cannot be used interchangeably and bias cannot be corrected by using a simple correction factor.

Finally, a total of 37 healthy young subjects and 100 general mixed ICU patients were recruited for the clinical validation where a three minutes VOT was performed. Literature suggests that fixed time thresholds could introduce variability in ReO_2_ changes.^123^ On the other hand, due to the slower deoxygenation rate observed with hDOS, achieving a consistent 40% StO_2_, as suggested,^123^ threshold upon cuff release was challenging. This device offers a precise automatized VOT protocol thereby reducing any additional variability due to operator.

Optical and hemodynamic properties reported are similar to those in Ref.^116^ for the *brachioradialis* muscle. The hDOS device successfully differentiated healthy microcirculation from impaired states in the general mixed ICU group, particularly in the baseline StO_2_, BFI and MMRO_2_. Also VOT-derived parameters showed differences in parameters related to microvascular reactivity such as ReO_2_, AUC StO_2_ and AUC BFI.

These findings align with literature,^29–31, 31–45, 124–126^ although studies often use the thenar muscle due to its accessibility.

The lack of standardized VOT protocols for NIRS technology is another significant concern. Variations in probe positioning, cuff size, inflation pressures, and VOT durations across studies complicate comparisons.^28, 30, 43, 49, 53, 57, 62, 71, 103, 123, 127–137^

On the other hand, when comparing our results specifically for the healthy population with what is present in the literature, a high variability is shown in the reported baseline values for StO_2_ in healthy subjects. In fact, baseline varies across devices generally ranging between 65% to 87%. Also DeO_2_, ReO_2_ and AUC StO_2_ differ widely in the reported healthy populations, which reflects both methodological differences (e.g. how the fitting point are chosen) and device sensitivities (e.g. source and detector distance in the probe, technology used, etc.) The hDOS TD-NIRS presented in this work for the healthy group, it shows slower DeO_2_ and ReO_2_ of what is normally reported. For example, ReO_2_ reported in the literature, ranges from ≈1.2 to ≈ 9.5 %/s. A comparison with AUC StO_2_ is more complex due to inconsistent reporting. In particular, the variability on ReO_2_ and AUC StO_2_ are due also to variations in protocols where VOT of 3 minutes or 5 minutes duration are often used.^30, 44, 73, 77, 130, 138–141^ These considerations stress the need of a standardized way of measuring the microcirculation in the healthy and ICU population.

The hDOS device demonstrates promising clinical applications. In critical care, it aids in assessing tissue perfusion and oxygenation, monitoring microcirculatory changes, and evaluating endothelial dysfunction, among the others.

## 5 Conclusion

The hDOS device is a hybrid diffuse optical platform that has been developed for application in the critical care, but given its performances it can find potential applications in many other fields, such as operating rooms and emergency care. The platform has been proven to be more accurate than a commercially available device.

## Data Availability

Data will be made available by the corresponding author upon reasonable request taking into account the appropriate norms for personal data privacy.

## Acknowledgments

This work has received funding from: the EuropeanUnion’s Horizon 2020 research and innovation programme under grant agreements No. 101016087 (VASCOVID), No. 101017113 (TINY-BRAINS), No. 871124 (LASERLABEUROPE V) and under the under the Marie Skłodowska- Curie grant agreement No. 101062306, Fundació CELLEX Barcelona, Fundació Mir-Puig, Agencia Estatal de Investigación (PHOTOMETABO, SCOSWEAR, SCOSDET), the “Severo Ochoa” Programme for Centres of Excellence in R&D (CEX2019-000910-S), LUX4MED, Generalitat de Catalunya (CERCA, AGAUR-2022-SGR-01457, RIS3CAT-001-P-001682 CECH), and Secretaria d’Universitats i Recerca del Departament d’Empresa.

## Appendix

### 5.1 Data quality

#### 5.1.1 TD-NIRS quality parameters

In Fig 17 a), the IRF/phantom box described in Section 2 is shown. If the probe is inserted with the optics facing upwards, the IRF is acquired; otherwise, a phantom measurement is obtained. To monitor the data quality of the TD-NIRS module, the IRF resolution, its shape, and temporal stability must be evaluated through its full-width-half maximum (FWHM) and barycenter. A summary of the quality parameters is presented in Fig 17 b). For clarity, only a portion of the temporal window where the distribution of time-of-flight is reconstructed is highlighted. The TD-NIRS laser is driven at 53 MHz corresponding to an available temporal window of approximately 19 ns wide and a timing resolution of 9.76 ps. For each measurement, one IRF is collected with an integration time of 1 s targeting 10^6^ counts per second per wavelength. In Fig 17 c), the results for the FWHM (in ps) and barycenter position in the temporal window (in ns) are reported, with shaded areas indicating the standard deviation calculated over seven months of measurements, for both 685 and 828 nm. For the barycenter here the mean ± standard deviation is reported, alongside the coefficient of variation (CV = standard deviation/mean). For the FWHM only the CV is reported. As mentioned in the main text, the barycenter of the DTOF at 685 nm was 3.9± 0.02 ns (CV = 0.4 %), and at 828 nm it was 3.6 ± 0.02 ns (CV = 0.6 %). The CV of the FWHM was 2.9 % for 685 nm and 1.1 % for 828 nm. In Fig. 18, the results obtained from fitting for *µ*_a_ and *µ*′_s_ at both 685 and 828 nm, based on the 31 phantom measurements collected in month 3, are reported. Each point in the distribution correspond to the single phantom repetitions (31 measurements x 20 repetitions). In Fig. 19, the effective total hemoglobin (tHb^ph^) and tissue oxygen saturation (StO_2_^ph^) extracted are shown. The distributions of the results, obtained by fitting the convolution of the DTOFs acquired with the corresponding IRF of the day (labeled “day” in the figure), were compared to those obtained by convoluting the DTOFs with the first IRF acquired in that specific month (labeled ”month” in the figure), using the Wilcoxon sign-rank test, with significance set at p*<*0.05. A larger CV was found in both *µ*_a_ (CV = 2.8%) and *µ*′_s_ (CV = 2.5%) for both wavelengths. This translated into a CV of 2.8% and 1.0% in tHb^ph^ and StO_2_^ph^. However, no statistically significant difference was found when comparing the “day” results to the“month” results for *µ*_a_ (p = 0.39 for 685 nm and p = 0.59 for 828 nm) and *µ*′_s_ (p = 0.15 for 685 nm and p = 0.19 for 828 nm).

**Fig 17.**
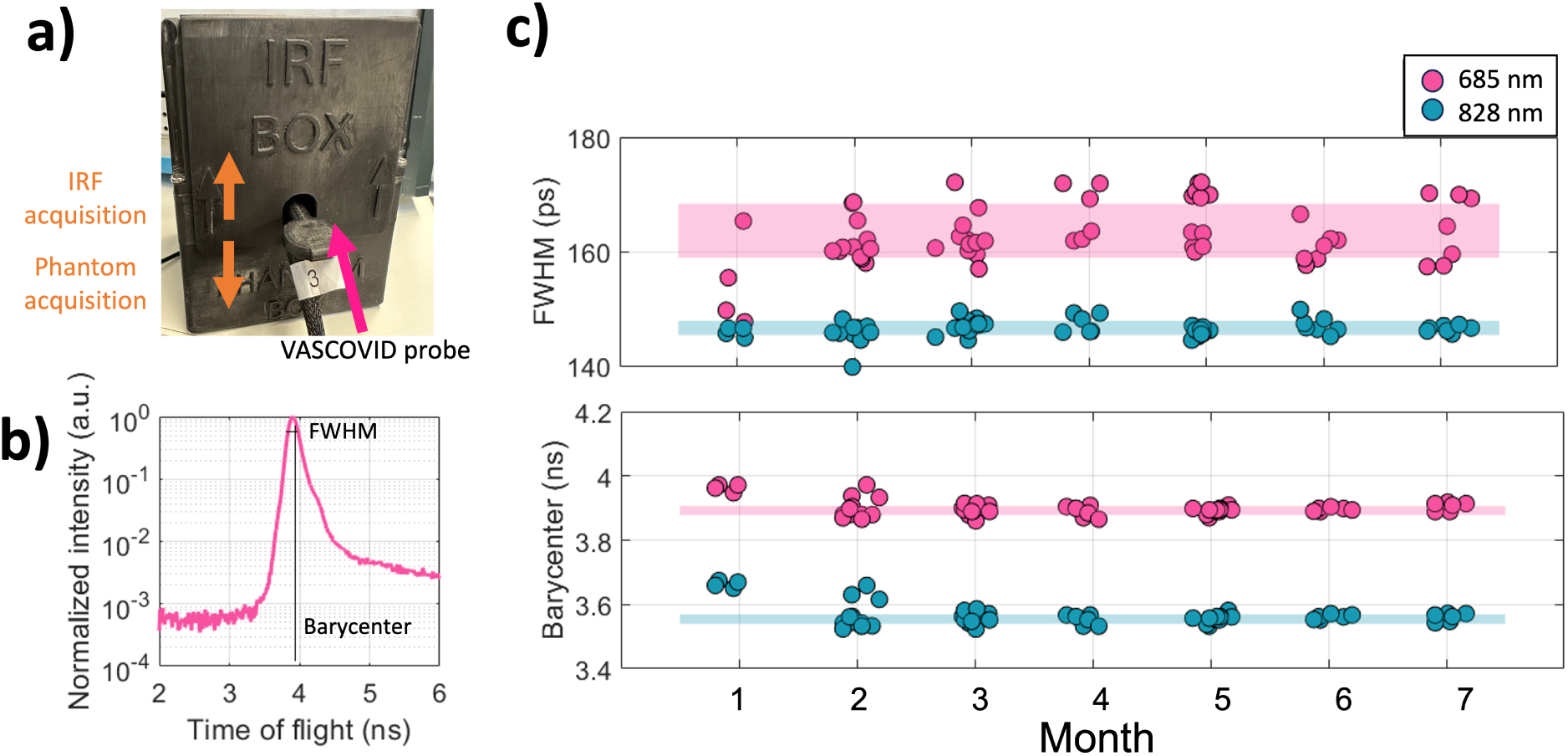
a) smart IRF and phantom box, with the probe optics facing upwards to obtain an IRF. b) An example of an IRF acquired at 685 nm is displayed, with the FWHM and barycenter figure of merit highlighted. c) The graph presents the FWHM and barycenter data for the 59 days of measurements, grouped by month for clarity. The shaded areas represent the standard deviation over all measurements.

**Fig 18.**
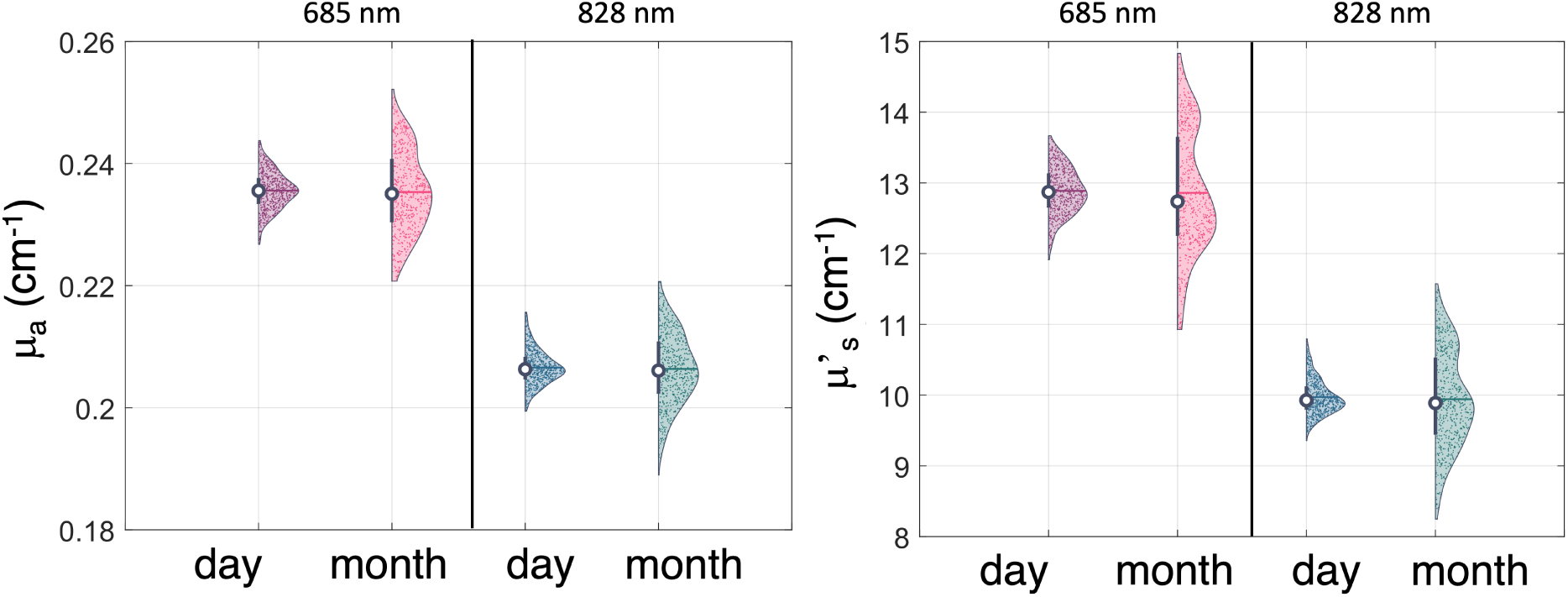
*µ*_a_ and *µ′*_s_ fitted at 685 and 828 nm for both the day-by-day (day) and month evaluation (month). Each dot in the distribution correspond to a single repetiton in each phantom measurement.

**Fig 19.**
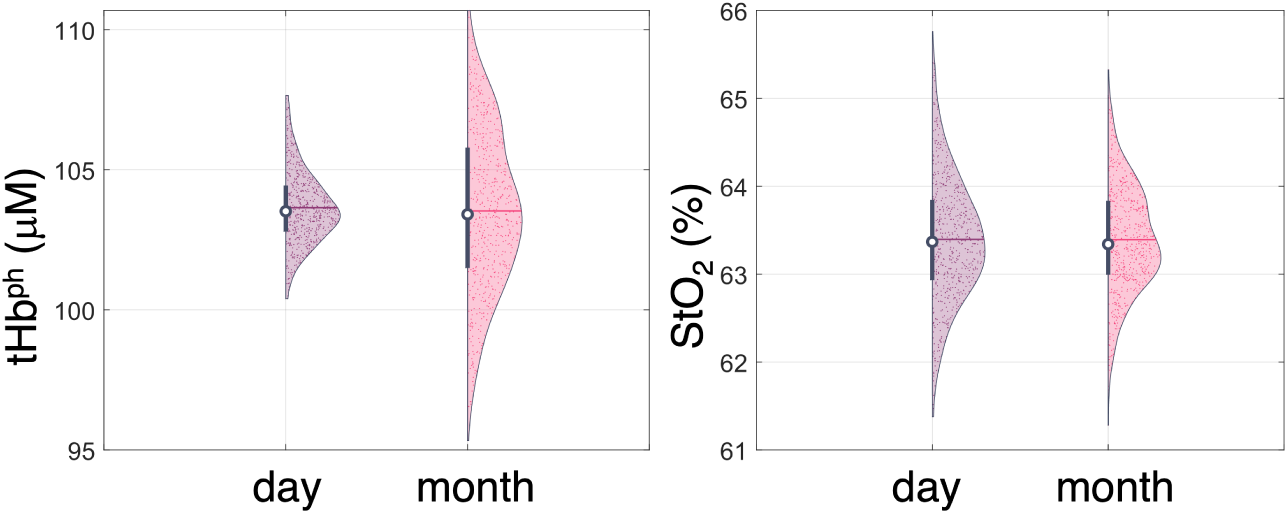
StO_2_^ph^ and tHb^ph^ for both the day-by-day (day) and month evaluation (month). Each dot in the distribution correspond to a single repetiton in each phantom measurement.

#### 5.1.2 DCS quality parameters

To assess the performance of the DCS, the count rate (in kHz) and the *β* parameters are evaluated. An example is depicted in Fig. 20, where the intensity of the detected DCS signal at the baseline of a healthy subject and the *β* parameter are shown. The *β* parameter, which depends on the number of modes detected, is calculated as the weighted average of the 2nd to the 4th bin of the autocorrelation function g_2_. In this example, a *β* value of 0.49±0.01 at the baseline is reported.

**Fig 20.**
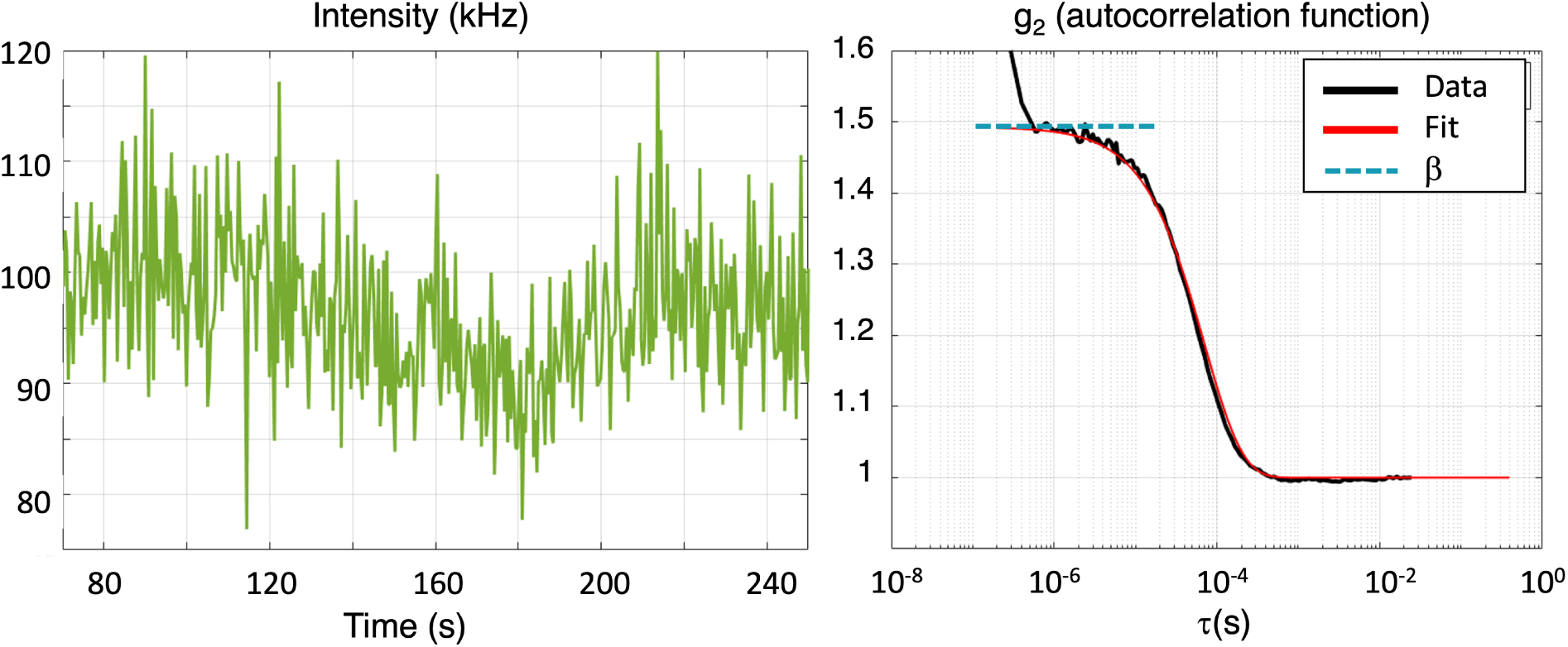
Left panel: intensity recorded during the baseline of a VOT protocol in an healthy subject. The blue shaded area represents the 10 s of integration time. Right panel: intensity autocorrelation function (g_2_) as a function of the lag time (*τ*), averaged over 10s of measurement. The *β* parameter is calculated as the weighted average of the 2nd to the 4th bin of the g_2_(*τ*).

### 5.2 Data and information saved

In this section, we report the list of all the variables that are recorded. Each module communicates with the single board computer (SBC) as described in Section 2 and the list of variables are listed and briefly described in Table 5 for the TD-NIRS module, in Table 6 for the DCS module, in Table 7 for the sensor and safety boards and finally in Table 8 for the PPG module.

**Table 5.** Data and information stored by the software from the TD-NIRS module. SBC: single board computer; DCS: diffuse correlation spectroscopy.

| Variable name | Units | Module origin | Frequency (Hz) | Description |
| --- | --- | --- | --- | --- |
| system time TD-NIRS | unix | SBC OS | 1 | time stamps SBC in the TD-NIRS reference frame |
| TD-NIRS time | ms | TD-NIRS | 1 | time stamps TD-NIRS module |
| DTOF 685 | photons | TD-NIRS | 1 | DTOF at 685 nm |
| count rate 685 | photons/s | TD-NIRS | 1 | count rate |
| $\mu_a$ 685 | 1/cm | TD-NIRS | 1 | real time absorption coeff. |
| $\mu'_s$ 685 | 1/cm | TD-NIRS | 1 | real time reduced scattering coeff. |
| peak position dtof 685 | - | TD-NIRS | 1 | bin corresponding to the peak value of the DTOF at 685 nm |
| chi2 685 | - | TD-NIRS | 1 | real time chi2 from fitting procedure (fit goodness) |
| dtof 685 fwhm | - | TD-NIRS | 1 | full width half maximum for the DTOF at 685 |
| DTOF 828 | photons | TD-NIRS | 1 | DTOF at 828 nm |
| count rate 828 | photons/s | TD-NIRS | 1 | count rate |
| $\mu_a$ 828 | 1/cm | TD-NIRS | 1 | real time absorption coeff. |
| $\mu'_s$ 828 | 1/cm | TD-NIRS | 1 | real time reduced scattering coeff. |
| peak position dtof 828 | - | TD-NIRS | 1 | bin corresponding to the peak value of the DTOF at 828 nm |
| chi2 828 | - | TD-NIRS | 1 | real time chi2 from fitting procedure (fit goodness) |
| dtof 828 fwhm | - | TD-NIRS | 1 | FWHM for the dtof at 828 nm |
| $\mu_a$ 785 | 1/cm | TD-NIRS | 1 | real time extrapolated absorption coeff. at 785 nm (See Section) |
| $\mu'_s$ 785 | 1/cm | TD-NIRS | 1 | real time extrapolated reduced scattering coeff. at 785 nm (See Section) |
| HbO | $\mu\text{M}$ | TD-NIRS | 1 | real time oxy hemoglobin |
| HbR | $\mu\text{M}$ | TD-NIRS | 1 | real time deoxy hemoglobin |
| tHb | $\mu\text{M}$ | TD-NIRS | 1 | real time total hemoglobin |
| StO<sub>2</sub> | % | TD-NIRS | 1 | real time tissue oxygen saturation |
| TOE | % | TD-NIRS | 1 | real time tissue oxygen extraction ($\text{SpO}_2\text{-StO}_2$) |
| MMRO<sub>2</sub> | %·cm<sup>2</sup>/s | TD-NIRS | 1 | real time metabolic rate of oxygen consumption ≈ TOE·BFI |

**Table 6.**
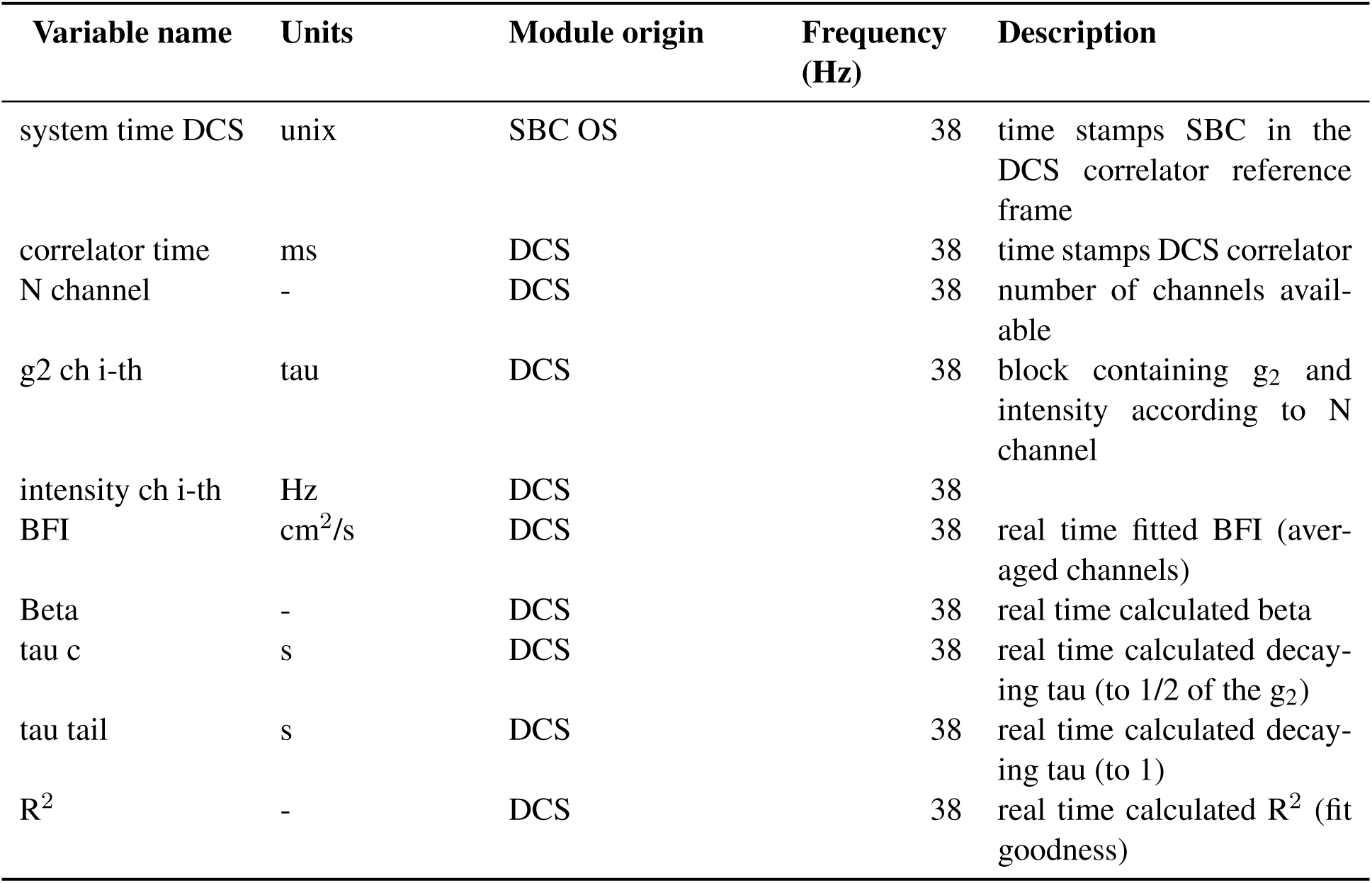
Data and information stored by the software from the DCS module. SBC: single board computer; DCS: diffuse correlation spectroscopy.

**Table 7.**
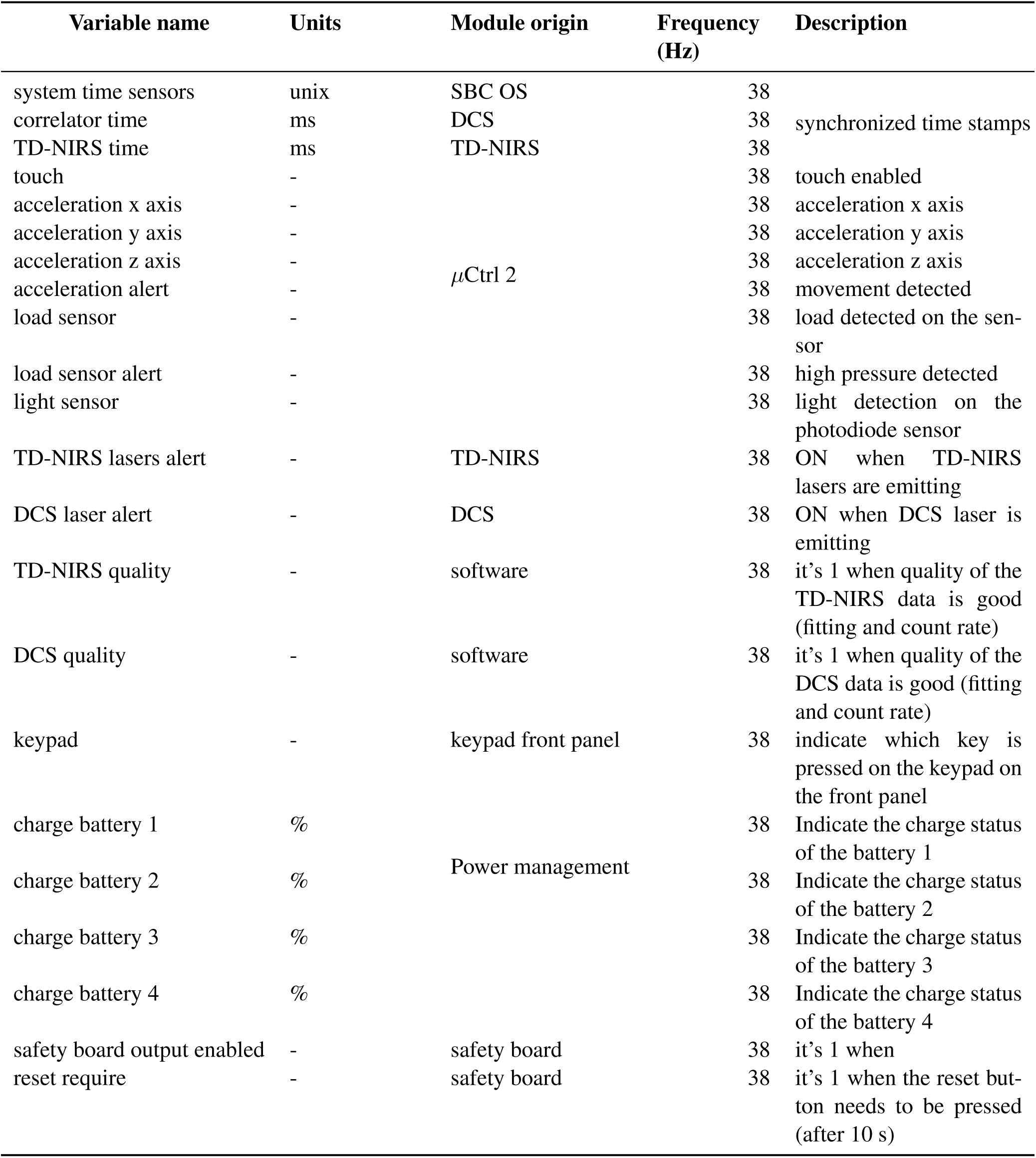
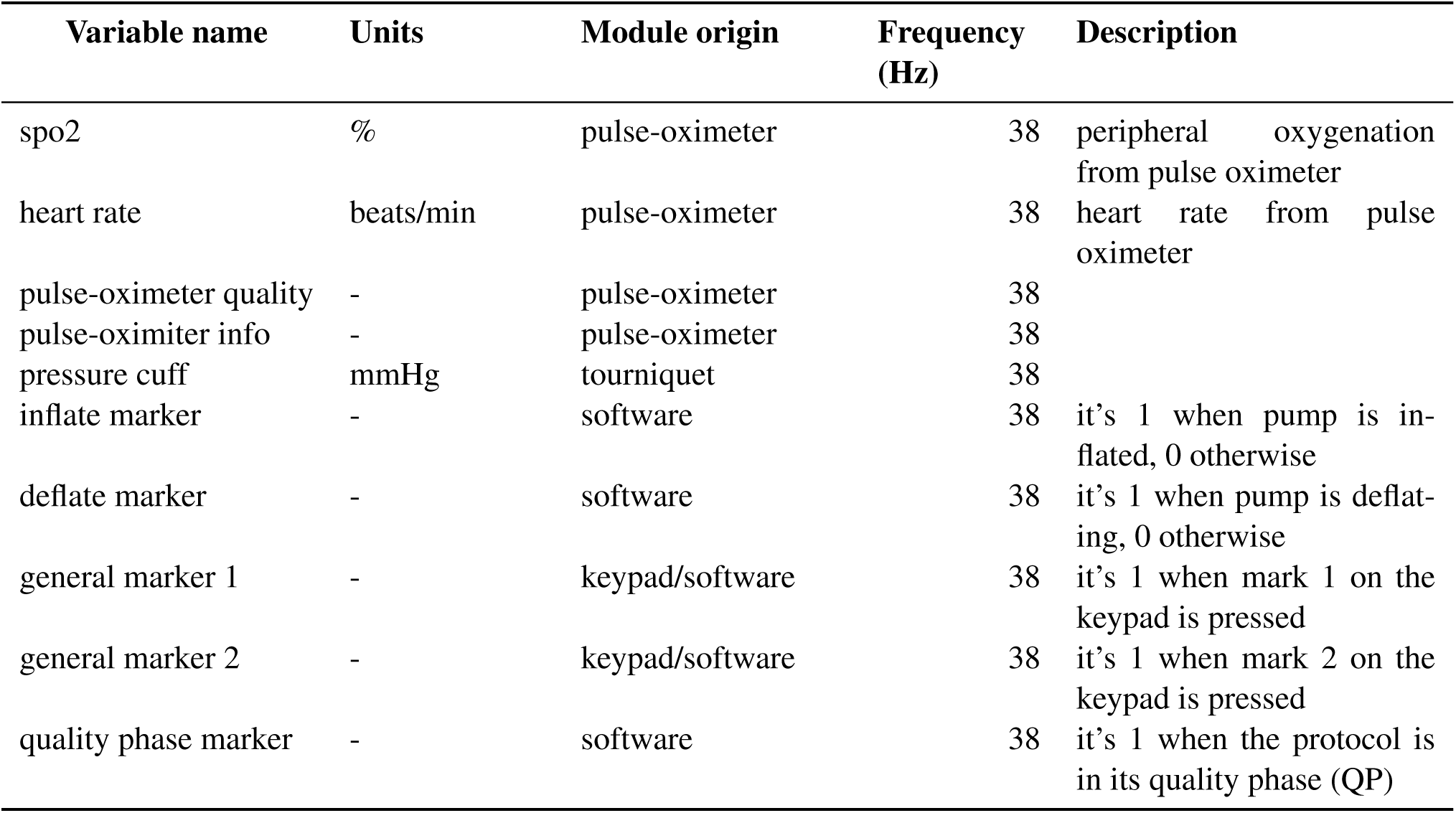
Data and information stored by the software from the various modules. SBC: single board computer; DCS: diffuse correlation spectroscopy; TD-NIRS: time-domain near-infrared spectroscopy.

**Table 8.**
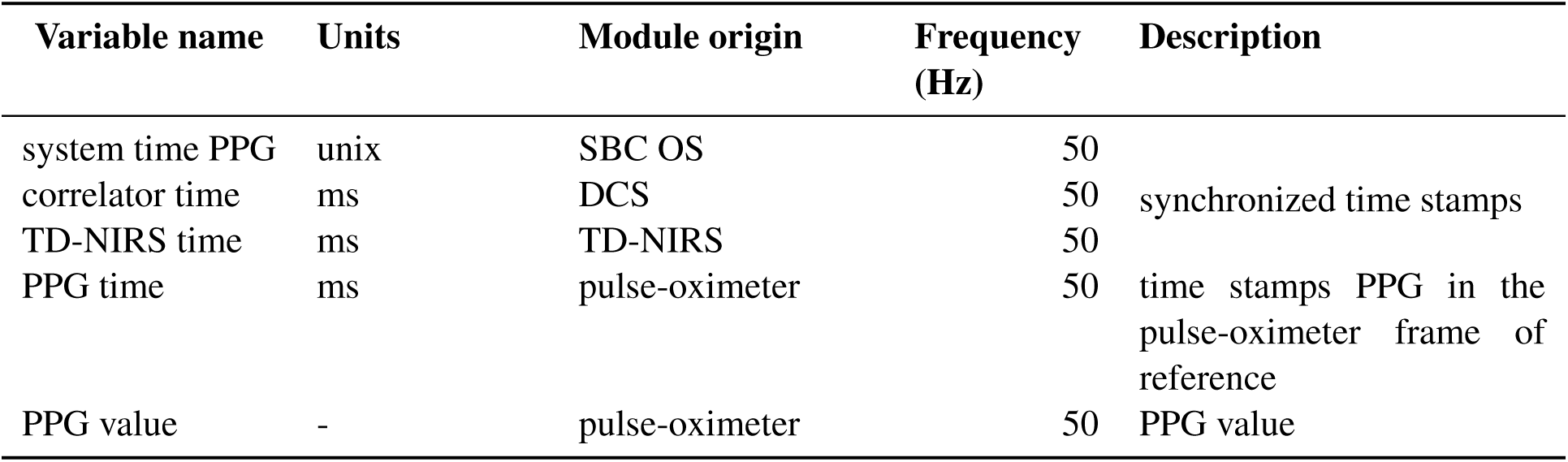
Data and information stored by the software from the SpO_2_ module. SBC: single board computer; DCS: diffuse correlation spectroscopy; TD-NIRS: time-domain near infrared spectroscopy.

## Footnotes

1 Developed during the VASCOVID project – European Union Horizon 2020 research and innovation, No. 101016087

2 We note that the current usage has reached 24 months with over 500 sessions on 410 patients under 12 protocols. We continue to monitor the quality and stability of the device and this will be reported along each particular clinical study results.

